# Impacts of the United Kingdom’s Soft Drinks Industry Levy: a systems-thinking informed systematic scoping review

**DOI:** 10.1101/2025.04.30.25326734

**Authors:** Catrin P. Jones, Jean Adams, Miriam Alvarado, Hannah Forde, Harry Rutter, Veronica Phillips, Roxanne Armstrong-Moore, Élisabeth Demers-Potvin, Adam Briggs, Steven Cummins, Oliver Mytton, Tarra L Penney, Mike Rayner, Nina Rogers, Peter Scarborough, Richard Smith, Martin White

**Author notes:** **Corresponding Author:** Martin White, MRC Epidemiology Unit, Level 3 Institute of Metabolic Science, University of Cambridge School of Clinical Medicine, Cambridge CB2 0SL, United Kingdom.

## Abstract

**Background:** Consumption of sugar-sweetened beverages (SSBs) is associated with weight, weight gain and incidence of a number of chronic diseases. The World Health Organization recommends taxation on SSBs to reduce consumption. In 2018 the United Kingdom introduced the Soft Drinks Industry Levy (SDIL), a tiered tax on manufacturers and importers of SSBs. We aimed to review the consequences of the SDIL across all potential outcomes, informed by a systems thinking approach, to understand the range and importance of its effects.

**Methods:** We undertook a systematic scoping review of empirical studies of the SDIL. We used a conceptual systems map of the hypothesised pathways of effect to inform data extraction and narrative synthesis. Findings are presented in an evidence map and their consistency assessed.

**Results:** 38 studies met our inclusion criteria. The SDIL was consistently associated with reformulation of soft drinks to reduce sugar content. It was also consistently associated with: reduced purchasing of sugar from eligible drinks without increasing purchasing of substitute products such as alcohol and confectionary; longer-term improvements in acute and chronic health outcomes; and reduced health and social care costs, with few negative economic impacts for industry.

**Conclusions:** By systematically mapping all outcomes evaluated, we have demonstrated the systemic and interconnected impacts of the SDIL. Further research should seek deeper understanding of how to evaluate such interventions as events in complex adaptive systems.

## 1. INTRODUCTION

Consumption of sugar-sweetened beverages (SSBs) is associated with adverse outcomes, including incidence of cardiovascular disease,^1^ type II diabetes,^1, 2^ obesity^3, 4^ and dental caries.^5^ More than 100 countries have implemented a tax targeting SSBs,^6^ and systematic reviews of their impact indicate that they encourage reformulation, increase prices and reduce sales of SSBs.^7, 8^

On 16^th^ March 2016, the United Kingdom (UK) government announced an excise tax on SSBs: the Soft Drinks Industry Levy (SDIL).^9^ Described as intended to “help with children’s health” by incentivising reformulation,^9^ the SDIL was a tax on manufacturers and importers, and a major policy within the government’s childhood obesity strategy.^10^ To allow industry reasonable time to prepare, a two-year period was planned between announcement and implementation in April 2018.^9^ The SDIL has two tiers – at implementation these were £0.18 per litre for drinks containing ≥5g and <8g sugar per 100mL and £0.24 per litre for drinks containing ≥ 8g sugar per 100mL. This tiered structured is consistent with World Health Organization (WHO) recommendations for SSB taxation in settings with high administrative capabilities.^11^ Drinks containing more than 75% milk and milk substitutes, pure fruit juices, alcohol replacements and other drinks are exempt from the SDIL, as are drinks produced by companies manufacturing <1 million litres/year.^12^

In their guidance on SSB taxation, the WHO states *“The best evaluations acknowledge contextual factors, feedback loops, the possibility of unintended consequences and the interplay of other external factors such as political support, public opinion, social norms and industry response (pp 319-321).“*^11^ This reflects a need for evaluations that explore how SSB taxes interact with the complex adaptive systems in which they are implemented. Complex adaptive systems are characterized by interconnected, interdependent elements that can adapt and evolve in response to changes in other parts of the system.^13^ Scholars have advocated for the use of systems thinking – which recognises these issues of complex adaptation - in evaluations,^14^ but previous systematic reviews of SSB taxes have tended to focus on one, or a limited number, of outcomes in insolation. These have included: changes in sales, purchasing, consumption, disease incidence, or public or political acceptability.^7, 8, 15–17^ Adopting a systems-thinking approach should allow assessments of SSB taxation to move beyond narrow questions concerning single, or closely related, outcomes to broader questions concerning the wider, intended, unintended and unanticipated, systemic impacts, as well as adaptations of the intervention and system to each other.^18^ This approach could both help with the framing of future evaluations and the future consideration of policy options.^14, 19^

When it was first implemented, the SDIL was relatively unusual in its design as a tiered levy targeting manufacturers and explicitly incentivising reformulation. ^11,12,23^ We conducted an evaluation of the SDIL informed by a systems perspective.^20^ Alongside this, further research was undertaken by others. No review has yet explored the full range of evidence on the impacts of the SDIL. In this research we therefore aimed to fill this gap by conducting a systematic scoping review of the impacts of the UK SDIL, identifying and synthesising findings across multiple outcomes using a systems-thinking informed approach.

## 2. METHODS

### 2.1. Study design

Given the heterogenous nature of the literature that we sought to synthesise, involving multiple methods, outcomes and study designs, a systematic scoping review was most appropriate.^21, 22^ The Joanna Briggs Institute Guidance^23^ and O’Malley’s six-step process^22^ informed the development of the methods. Our review adhered to the Preferred Reporting Items for Systematic Review and Meta-Analysis – Extension for Scoping Reviews (PRISMA-ScR)^24^ and the protocol was published.^25^ In keeping with systematic scoping review methodology, we did not examine effect sizes nor assess risk of bias.^24^

### 2.2. Theoretical framework

A conceptual systems map, developed to underpin our evaluation of the SDIL,^20^ underpinned our synthesis (Figure 1). Developed in an expert consensus workshop and refined using a Delphi study and interviews with stakeholders,^26^ the map theorised the potential impacts of the SDIL across the system in which it was introduced. We refer to the variables in the map as ‘nodes’ and the hypothesised pathways between variables as ‘connections’, and use these terms throughout as appropriate.

**Figure 1:**
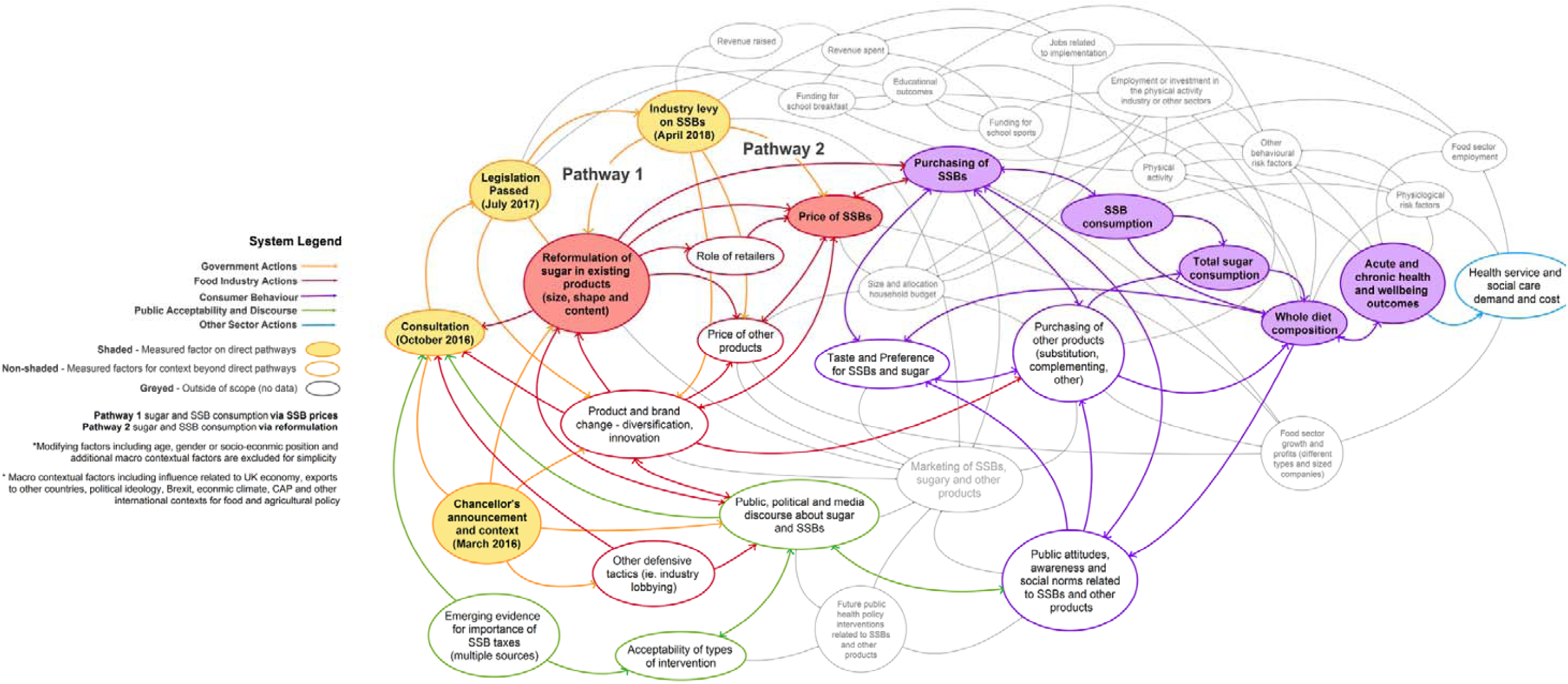
Conceptual system map developed to inform the (NIHR-funded) evaluation of the SDIL^20^

### 2.3. Information sources

We searched the following databases of peer reviewed literature in March 2024: Medline (via Ovid), Embase (via Ovid), Web of Science (core collection), Global Health (via Ebscohost), EconLit (via Ebscohost), ERIC (via Ebscohost), Sociological Abstracts (via ProQuest), PsycInfo (via Ebscohost), and Worldwide Political Sciences Abstracts (via ProQuest). Grey literature sources were searched up to November 2022 including government websites, non-government organisation (NGO) websites, preprint servers, Ethos, trial databases, UK funders’ websites, and targeted searches of google.co.uk. A full list of grey literature sources is shown in Appendix A. Corresponding authors of included studies found via searches were contacted to seek additional relevant unpublished work.

### 2.4. Search strategy

The search strategy was developed through initial scoping searches and with support from an academic librarian (VP). Table 1 shows the logic grid used to underpin the searches of peer reviewed literature, which was adapted for each source. Exact search terms for databases of peer reviewed literature are shown in Appendix B and those for grey literature in Appendix A.

**Table 1:**
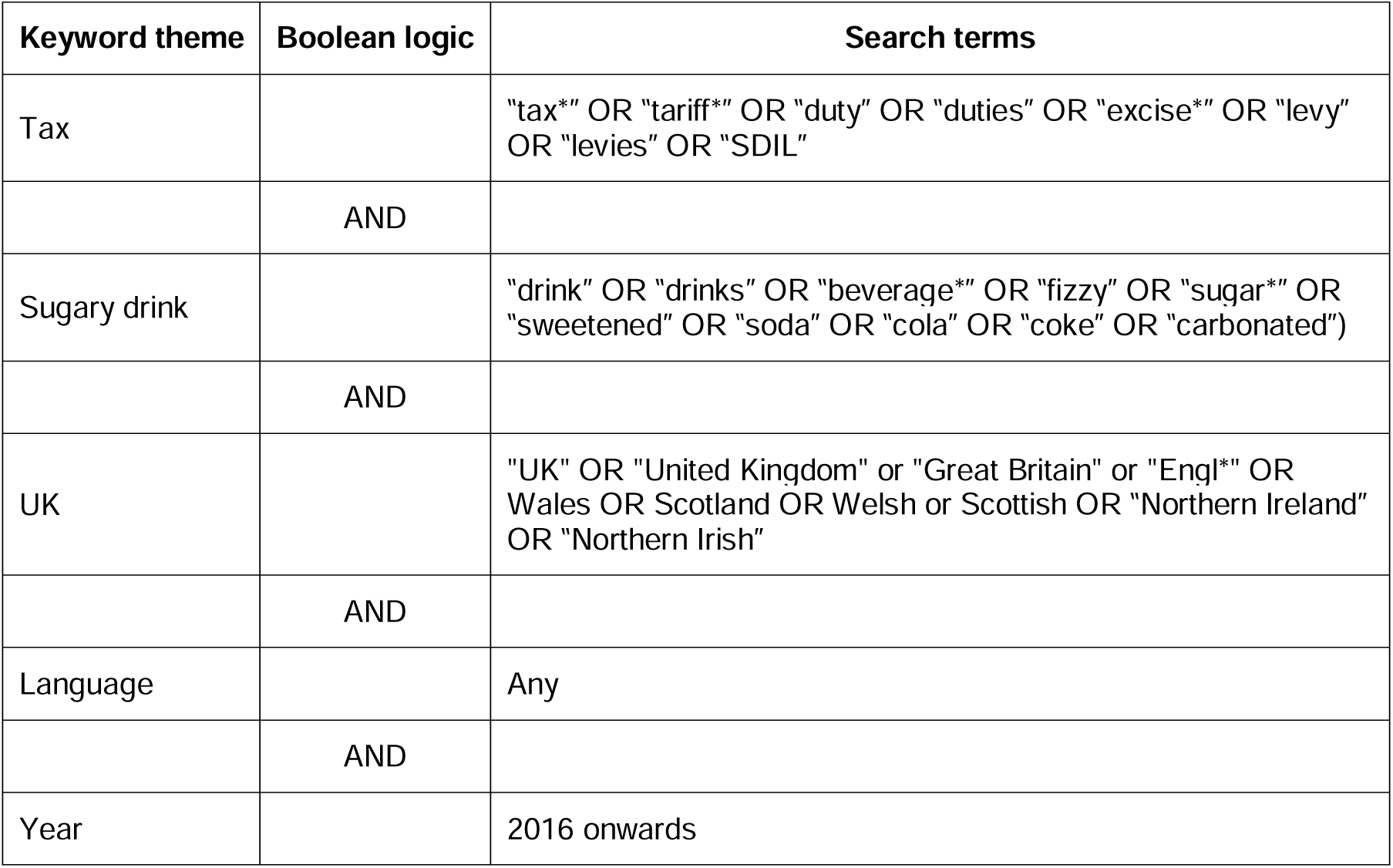
Logic grid for searching databases.

| Keyword theme | Boolean logic | Search terms |
| --- | --- | --- |
| Tax |  | "tax*" OR "tariff*" OR "duty" OR "duties" OR "excise*" OR "levy" OR "levies" OR "SDIL" |
|  | AND |  |
| Sugary drink |  | "drink" OR "drinks" OR "beverage*" OR "fizzy" OR "sugar*" OR "sweetened" OR "soda" OR "cola" OR "coke" OR "carbonated") |
|  | AND |  |
| UK |  | "UK" OR "United Kingdom" or "Great Britain" or "Engl*" OR Wales OR Scotland OR Welsh or Scottish OR "Northern Ireland" OR "Northern Irish" |
|  | AND |  |
| Language |  | Any |
|  | AND |  |
| Year |  | 2016 onwards |

### 2.5. Eligibility criteria

We sought evidence on connections between nodes representing the SDIL on the one hand and relevant proximal or distal outcomes across any domain on the other, published on or after 16^th^ March 2016. An inclusive approach was taken with few restrictions on study design, methods, population, or outcome. Table 2 shows the full inclusion and exclusion criteria.

**Table 2:** Inclusion and exclusion criteria.

|  | **Inclusion criteria** | **Exclusion criteria** |
| --- | --- | --- |
| Date of publication | From 16<sup>th</sup> March 2016 (inclusive) | Before 16<sup>th</sup> March 2016 |
| Publication type | Articles, books, book chapters, data papers, short communications, meeting abstracts, meeting summaries, proceedings papers or reports, published by any organisation | Documents only expressing opinions: e.g., blogs, editorials, commentaries |
| Intervention/ Exposure | Primary focus on the UK Soft Drinks Industry Levy applied to sugary beverages in the UK from 1<sup>st</sup> April 2018 (referred to as the sugar tax, SDIL, etc.) | Focus on tax, levy or duty applied to other commodities in the UK e.g., alcohol, tobacco; other fiscal measures applied to SSBs in the UK (e.g. subnational taxes or hypothetical taxes); or fiscal measures applied to sugary beverages elsewhere |
| Study type or method | Studies that analyse data related to SDIL outcomes, collected using any method. | No data analysed |
| Country | Study of the SDIL in the UK context | Not UK context |
| Language | Any | None |
| Outcomes | All (including e.g. dietary or health outcomes, changes in products economic factors, attitudes or behaviours etc.) assessed in relation to period after the SDIL was announced on 16<sup>th</sup> March 2016 | None |

### 2.6. Deduplication and screening

Results from searches of databases of peer-reviewed literature were deduplicated using Endnote X20 (Clarivate), and remaining results were uploaded to Covidence (Veritas Health Innovation Ltd) together with the grey literature results, where a second deduplication was undertaken. Titles and abstracts (or equivalent) were independently screened in duplicate (by authors MA, CPJ, HF) against the eligibility criteria listed in Table 2. Screeners met to discuss and refine the eligibility criteria. Full texts were retrieved for studies retained after title and abstract screening

### 2.7. Data charting and synthesis

Data extraction forms were piloted on a sub-sample of included texts by CPJ and MA and then finalised after discussion. Data was extracted into Microsoft Excel by CPJ, MA, EDP and RAM, who compared extraction spreadsheets and met to discuss any discrepancies. Data was extracted from study abstracts rather than full texts, although full texts were explored if details were unclear in abstracts. This approach was chosen after extensive piloting to ensure author-determined ‘key findings’ were selected for synthesis, representing the most important findings and avoiding an overly complex analysis.

To generate an initial list of relevant nodes, we first extracted those represented on the SDIL conceptual system map.^20^ As far as possible, each variable identified from an included study was then allocated to the most relevant node in this list, following the clustering approach used by Alvarado *et al* (2023).^18^ Relevant variables identified in the included texts that did not represent a good match with nodes in our initial list were added to the list as new nodes (see full list in appendix D).

Connections between nodes were derived by extracting hypothesised pathways between two variables identified in each study abstract. As a check on the reliability of extraction of connections from abstracts, a sample of six studies were scrutinized by JA, RAM, HR and MW, with each study read in full by two authors who independently extracted all connections identified, with results compared to those previously extracted from abstracts only.

Connections between nodes were assigned a direction (i.e. from node A to node B) and polarity (positive, negative or neutral, where: positive polarity represents a change in the magnitude of the proximal variable leading to a change in magnitude of the distal variable in the *same* direction; negative polarity represents a change in the magnitude of the proximal variable leading to a change in magnitude of the distal variable in the *opposite* direction; and neutral represents a relationship with no clear polarity). Judgements concerning direction and polarity of connections were based on the consensus views on those extracting data after close reading of included texts.

In a number of cases, connections between the same pairs of nodes were reported in more than one study. In these cases, we estimated consistency of connections as follows: where at least two studies reported on the same connection, if at least 75% reported the same polarity, we designated this connection as ‘high consistency’; otherwise, connections were designated ‘low consistency’. Consistency was not estimated for connections reported by only one study. For example, for the connection between the SDIL and Purchases of SDIL ineligible drinks, there were five positive connections, one connection with no clear polarity and zero negative connections. This connection was designated ‘high consistency’ because there were six studies reporting on it and as at least 75% of the connections had the same polarity (i.e. five of six or 83%). In the case of the connection between the SDIL and measured negative health or wellbeing outcomes, there were zero positive connections, three connections with no clear polarity and three negative connections. This connection was designated ‘low consistency’, as less than 75% of connections (i.e. three of six of 50%) had the same polarity. To structure reporting, we grouped connections into eight thematic categories based on destination nodes.

We used Kumu (www.kumu-io)^34^ to visually display nodes and connections as well as the direction and polarity of connections.

### 2.8. Changes to protocol

We originally specified that we would assess the quality of studies.^25^ We piloted the use of both the Quality Assessment for Diverse Studies (QuADS)^27^ tool for academic literature and AACODS (Authority, Accuracy, Coverage, Objectivity, Date, Significance)^28^ tool for grey literature. Given the heterogenous nature of included studies, neither of these tools were suitable. We thus did not perform quality assessment. It is usual in scoping reviews not to include an assessment of study quality.^24^ Further changes to protocol were the inclusion of multiple researchers during data extraction rather than two; and the extension of narrative synthesis to include the visualisation of the findings using a systems thinking informed evidence map.

## 3. RESULTS

### 3.1. Selection and characteristics of sources of evidence

After duplicates were removed, 850 records were identified for screening, 183 full texts were reviewed and 38 studies met the criteria for inclusion in the review (Figure 2). A list of all included studies is in Appendix C. Seventeen lead authors of included studies were contacted, ten responded and two additional studies were included in the review from these contacts. Thirty-three studies were quantitative, two mixed-methods and three qualitative. Sixteen studies from the NIHR-funded SDIL evaluation were included.

**Figure 2:**
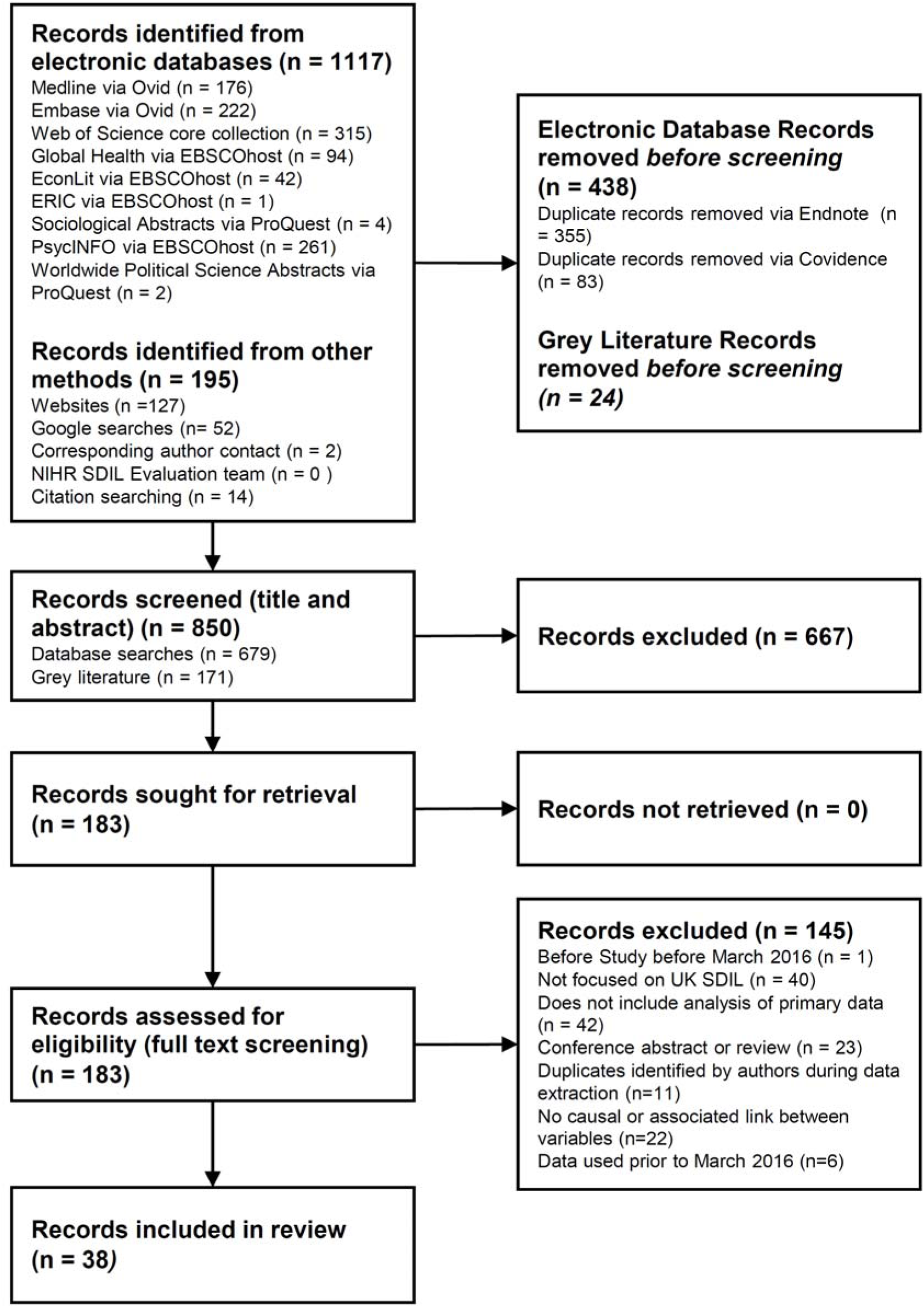
PRISMA 2020 flow diagram summarising searching and screening

### 3.2. Variables and nodes

From the 38 studies included we identified 265 variables, which were assigned to 41 unique nodes. A full list of all variables extracted from included studies and their allocation to nodes is shown in Appendix D.

Although some studies analysed data specifically related to the announcement (16^th^ March 2016) or implementation (5^th^ April 2018) of the SDIL, many did not explicitly analyse the impacts of either the announcement or implementation. We therefore included three categories of timepoint as nodes: announcement, implementation, and announcement and/or implementation (often stated in studies simply as ‘the SDIL’).

### 3.3. Direction, polarity, nature and consistency of connections

Appendix E lists all connections identified by frequency and identifies the studies from which they were extracted. In Appendix F, the polarity of all connections is presented with consistency designations. A visual representation of nodes and connections is presented in the evidence map in Figure 3. In this, lines represent the connections between nodes.

**Figure 3:**
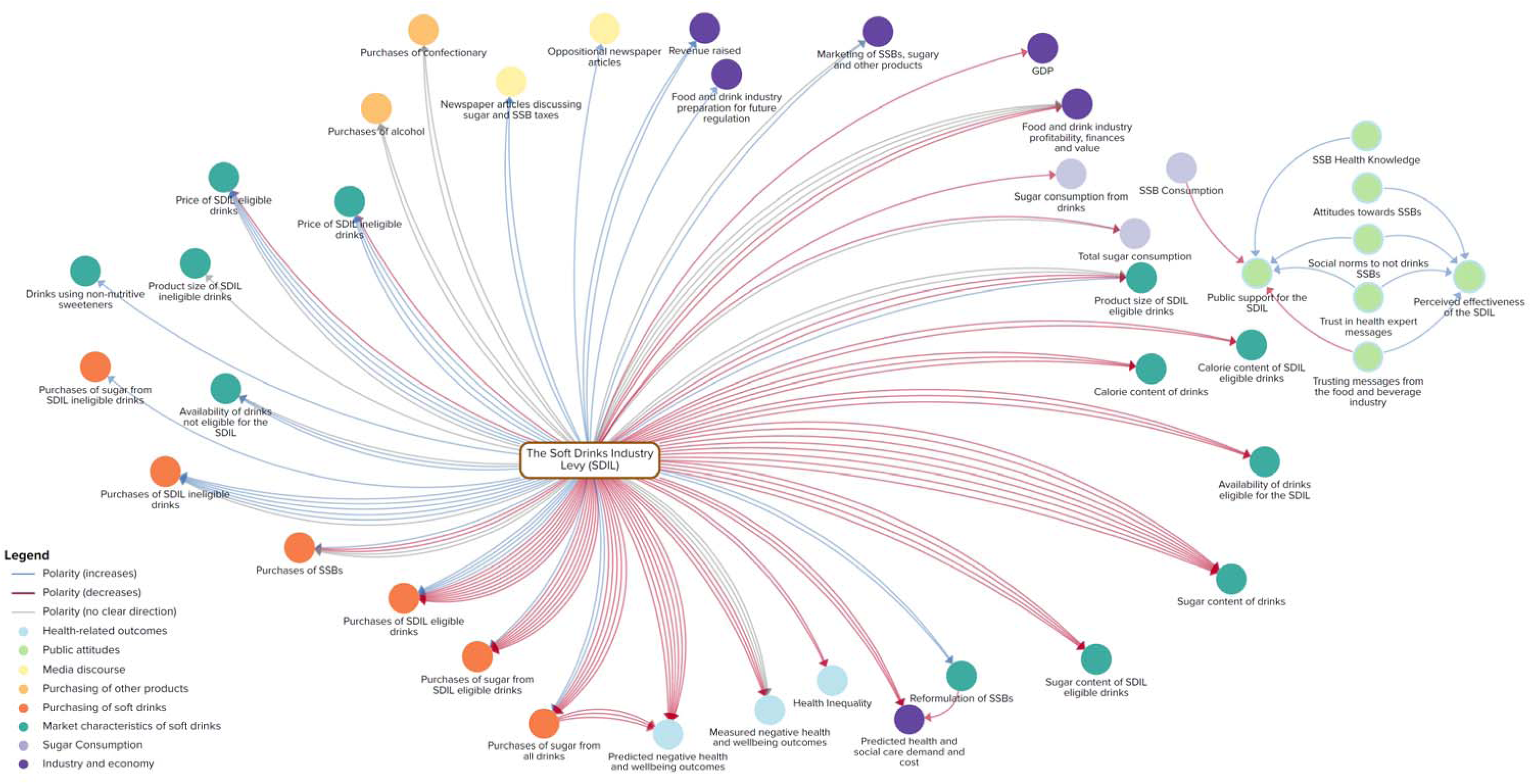
Map of nodes and connections identified in the review.

Connection direction is represented with arrows and polarity with colour (blue = positive polarity; red = negative polarity; grey = no clear polarity).

Overall, we identified 133 connections between 41 nodes in the 38 included studies. However, many of these connections were represented in more than one study, such that there were only 43 unique connections. We grouped connections in Figure 3 into 8 thematic categories, colour coded for visual clarity, as follows: market characteristics of soft drinks; media discourse; public attitudes; purchasing of soft drinks; purchasing of other products; sugar consumption; health-related outcomes; and industry and economy outcomes.

There were many more nodes and connections than we theorised in our conceptual system map. New nodes were particularly related to: soft drink industry actions and economic factors, broader economic impacts, and health inequality. Otherwise, new nodes primarily expanded and specified existing nodes in greater detail (e.g. dividing purchasing of sugar into purchasing from SDIL eligible and ineligible drinks; or expressing reformulation in terms of volumes of drinks, calorie content, sugar content and availability).

The weight of evidence for many connections is striking. Of 43 unique connections, 27 were reported by at least two studies. One unique connection was reported in 12 studies – that between the SDIL and purchases of SDIL eligible drinks. Other frequently reported unique connections were between the SDIL and: sugar content of drinks (n=8), purchases of sugar from SDIL eligible drinks (n=8) and all drinks (n=7), and predicted negative health and wellbeing outcomes (n=7).

There was high consistency of unique connections. Of 27 unique connections that were reported by more than one study, 17 (63%) were designated as ‘high consistency’ and 10 (37%) as ‘low consistency.

### 3.4. Connections designated to have high consistency

Table 3 summarises the number of unique connections, individual connections and consistency of individual connections within unique connections by category of destination node.

**Table 3.** Summary of connections by category of destination node.

| **Category of destination node** | **N unique connections** | **N individual connections** | **N (%) high consistency unique connections** |
| --- | --- | --- | --- |
| Market characteristics of soft drinks | 11 | 42 | 7 (64) |
| Public attitudes, awareness | 9 | 9 | 0 |
| Industry & economy | 7 | 15 | 2 (29) |
| Purchasing of soft drinks | 6 | 39 | 2 (33) |
| Health related outcomes | 4 | 18 | 3 (75) |
| Media discourse | 2 | 3 | 1 (50) |
| Purchasing of other products | 2 | 4 | 2 (100) |
| Sugar consumption | 2 | 3 | 1 (50) |

Eleven unique connections, including 42 individual connections, had a destination node categorised as ‘market characteristics of soft drinks’. Destination nodes in this group related to price, availability, product size and content of different groups of soft drinks. Seven of these 11 (64%) unique connections were designated ‘high consistency’. These seven unique connections indicated the SDIL was associated with decreased availability of drinks eligible for the SDIL, reduced calorie content of all drinks and SDIL eligible drinks, increased price of SDIL ineligible drinks, reformulation of SSBs, and reduced sugar content of drinks and SDIL eligible drinks.

Destination nodes were categorised as ‘public attitudes and awareness’ in the case of nine unique connections – which explored putative influences on perceived effectiveness of, and public support for, the SDIL. Each of these nine connections was included only once meaning that consistency could not be designated.

Seven unique connections, representing 15 individual connections, were categorised as having a destination node in the ‘industry and economy’ group. Two of these connections (29%) were considered to have high consistency. These indicated that the SDIL was associated with reduced predicted demand for, and cost of, health and social care services, and increased revenue raised.

A total of 39 individual connections within six unique connections had a destination node categorised as ‘purchasing of soft drinks’. Only two (33%) of these unique connections were designated as having high consistency indicating that the SDIL was associated with increased purchases of SDIL ineligible drinks and decreased purchased of sugar from SDIL eligible drinks.

Four unique connections containing 18 individual connections had a destinating node within the ‘health related outcomes’ group. Of these four, three (75%) unique connections were considered to have high consistency. These indicated that the SDIL was associated with reduced health inequality and predicted negative health and wellbeing outcomes. Purchases of sugar from drinks was also consistently reported to be associated with predicted negative health and wellbeing outcomes.

The destination node of two unique connections was grouped as ‘media discourse’. Only one (50%) of these was designated to have high consistency indicating that the SDIL was associated with an increase in newspaper article discussing sugar and SSB taxes.

Two unique, and four individual, connections had a destination node within the ‘purchasing of other products’ group. Both individual connections were considered to have high consistency. These found that the SDIL was associated with no change in purchases of alcohol and confectionary.

Sugar consumption was the destination node group for two unique connections representing three individual connections. Neither of these unique connections were considered to have high consistency.

## 4. DISCUSSION

### 4.1. Summary of main findings

This systematic scoping review of impacts of the UK SDIL, informed by a systems perspective, identified 38 studies assessing 43 unique connections between 41 nodes. A conceptual system map of SDIL impacts, developed by the authors before the SDIL was implemented, was used to inform analysis and develop a map of the evidence. For those 27 unique connections that were reported on two or more occasions, the polarity of association was consistent in around two-thirds (n=17) of occasions. Almost three quarters of unique connections (n=32, 74%) had an origin node of the SDIL making it difficult to operationalise chains of causation with more than two nodes.

Our findings suggest that the SDIL is likely to have achieved its aim to incentivise manufacturers to reformulate soft drinks to reduce sugar content. In particular, we found consistent evidence that the SDIL was associated with reduced availability of drinks eligible for the SDIL, reduced calorie and sugar content of all drinks and SDIL eligible drinks and increased reformulation of SSBs. We also found consistent evidence that the SDIL was associated with reduced purchasing of sugar from SDIL eligible drinks. These benefits occurred alongside no changes in purchases of alcohol or confectionary. Modelling studies found consistent associations between the SDIL and decreases in negative health and wellbeing outcomes. There was also consistent evidence that the SDIL had benefits for the economy with decreases in predicted demand for and cost of health and social care services, and increased revenue raised. Other associations were not considered consistent or were not investigated enough for us to designate an indicator of consistency.

### 4.2. Strengths and limitations

The use of a systems thinking approach and broad inclusion criteria ensured that we included the full range of intended, unintended and unexpected health, social and economic outcomes of the SDIL that have been studied over the short and medium term. Given the large number of potential impacts, the study benefited from a visualisation of the evidence base to illustrate polarity of possible impacts and consistency of these. We aimed to bring together all available evidence examining impacts of the SDIL, resulting in a large but highly heterogeneous dataset with had diverse measures, definitions, and parameters. Our approach prioritised breadth over depth and the use of scoping review methods precluded a quantitative synthesis of the SDIL’s impact that might have further advanced understanding of its systemic effects; this could be explored in future research. Applying an assessment of consistency informed our understanding of how the SDIL has been evaluated, and the conclusions we might draw. However, the threshold we set for consistency of polarity was arbitrary. Furthermore, whilst some connections were reported a number of times (e.g. 12 unique connections included five or more individual connections), 16 connections were only reported once meaning we did not designate a consistency category. A further potential limitation is the extraction of data from study abstracts only. Analysis in a subsample of studies comparing extracted connections from abstracts to those in the full text confirmed abstracts were a valid representation of full texts. Whilst we expect that we are likely to have identified all unique connections present in included studies, we may have missed additional individual connections.

It is possible, therefore, that some connections might be supported by more evidence than is currently present in the evidence map. It is also possible that there was some degree of ‘double counting’ with some studies presenting updates on previous ones (e.g. an analysis of impacts one year after implementation, followed by a further analysis two years after implementation)^29, 30^ reporting on similar analyses of the same dataset.

Additional connections between distal nodes may also exist in literature not specific to the SDIL, as well as within the full text of some studies reviewed in this study. Including an interrogation of every possible unique connection between all included nodes would have required a specific search strategy for each connection and likely would have resulted in an unmanageable dataset. Lastly, our visual evidence map does not reflect sub-populations and thus is limited in its ability to represent equity considerations.

### 4.3. What the study adds to prior knowledge

There have been few reviews that have summarised the broad range of outcomes of SSB taxes. For example, the most recent systematic review of SSB taxes focused on economic outcomes only (tax pass-through rate for prices, percentage reduction in SSB demand, and price elasticity of demand for sales and consumption).^7^ That review quantitatively summarized empirical evidence globally with a meta-analysis, but did not adopt a systems perspective. In contrast, we have reviewed evidence focused on one country and one tax regime but considered an extensive range of outcomes and interrogated the evidence terms of the direction and polarity of relationships, viewed through a complexity lens.

In contrast, Alvarado et al (2023), reviewed the global empirical literature on SSB taxes to identify the extent to which systems thinking was incorporated in studies, and document the influences on and impacts of SSB taxes explored in this body of knowledge.^18^ That review found that the majority of studies had focused on economic and health outcomes of SSB taxes, with fewer studies exploring broader societal outcomes such as economic growth, industry costs, or political factors. Only 3 out of 327 studies analysed by Alvarado et al (2023) explicitly adopted a systems thinking approach. Nevertheless, the authors concluded that most studies had the potential to shed light on the systemic impacts of SSB taxes.

In our study, by considering a wide range of factors in one setting, we have been able to develop a more cohesive narrative illustrating the complex relationships between the SDIL and a broad range of outcomes. Synthesising the totality of the evidence, both quantitative and qualitative, relating to this case study enables a deeper understanding of its systemic impacts. Nevertheless, the predominant approach of planning, conducting and publishing analyses of single pathways in each study, even if the broader intention is to explore the broad systemic effects of an intervention,^20^ makes develop an integrated systemic theory of change challenging.

### 4.4. Interpretation and implications for policymakers

Observations and reflections on the way public health policy is evaluated can be made based on these findings. Our conceptual systems map contained connections between nodes to depict and explain the complexity of the systemic impacts that could result from the SDIL.^20^ However, a key observation is that research to date has been concentrated on a relatively small number of direct and proximal pathways from the SDIL. For example, in the conceptual system map, interrelations between changes in formulation, purchasing and price were hypothesised,^20^ but we found they were not articulated in this way in the studies included in the review. Included studies simply studied the association between the SDIL and each outcome separately, rather than trying to disentangle any inter-relationships.

A second observation is that many studies investigated the same pathways, resulting in substantial weight of evidence for some connections and relatively little for others. Connections for which single studies provided evidence were either complex and challenging to conduct (e.g. the GDP node),^31^ on topic areas where qualitative interrogation is more common (e.g. the Food and drink industry preparation for future regulation node^32^), or perhaps on topics considered of less public health ‘importance’ (e.g. oppositional newspaper articles).^33^ Whilst having multiple consistent connections, as demonstrated in this review, provides substantial evidence of the impacts of the SDIL across a range of relevant public health outcomes, bias towards certain study types (often used repeatedly for the same question),^34^ limitations of existing hierarchies of evidence^35^ and bias towards asking narrow research questions^36^ could be limiting investigation into other systemic policy impacts.

### 4.5. Unanswered questions and future research

In this study we have identified a broad range of potential impacts of the SDIL, informed by a systems thinking approach. However, we have not further elaborated the conceptual system map,^20^ which acted as the starting point for this research, into a more detailed systemic theory of change. Doing so would enable further interrogation of the system in which the SDIL exerted its impacts, for example by identifying further leverage points for public health benefit, such as those represented by reinforcing feedback loops or exemplifying system archetypes.^13^ This represents a substantial body of work that could be the focus of a future study, perhaps by building on the methods of Alvarado et al (2023).^18^

More broadly, the idea of systems thinking informed evaluation is growing in popularity,^37^ but there remain few examples of studies that have managed to take a truly systemic perspective in evaluating a population level intervention. Further development of the field beyond inspiration by systems thinking to exemplification of systemic evaluation is now needed.

## 5. CONCLUSIONS

In this review, we have surfaced a high level of consistency in some findings of studies exploring the UK SDIL and important outcomes for public health. Overall, these suggest that the SDIL was consistently found to achieve its primary aim of reformulation of soft drinks to reduce sugar content. It also consistently reported to have resulted in reduced purchasing of sugar from eligible drinks without increasing purchasing of substitute products such as alcohol and confectionary, and in longer-term improvements in acute and chronic health outcomes, and reduced health and social care costs, with few long-term negative impacts for industry. Studies tended to focus on simple two-node pathways with few explicitly taking a systems approach. To better understand how population health interventions interact with the systems in which they are implemented, researchers and funders should look to move away from examination of a narrow set of variables and outcomes embedded in simple, linear pathways, and towards broader assessments of multiple connections between outcomes and their relationships informed by systems thinking.

## Data Availability

All data produced in the present study are available upon reasonable request to the authors

## Acknowledgements

For the purpose of Open Access the author has applied a Creative Commons Attribution (CC BY) licence to any Author Accepted Manuscript version arising. The authors thank the corresponding authors of included manuscripts for their time in responding to us and inclusion of work in progress in our review. The authors also thank other members of the SDIL Evaluation team, who contributed to the our evaluation and helped to inspire this review: Lauren Bandy, Linda Cobiac, Laura Cornelsen, Richard Harrington, Marcus R. Keogh-Brown, Cherry Law, David Pell, Henning Tarp Jensen, Dolly R. Z. van Tulleken.

## Data statement

The lead author has full access to the data reported in the manuscript.

