## Supplementary Appendices for "Impacts of the United Kingdom’s Soft Drinks Industry Levy: a systems-thinking informed systematic scoping review"

***Appendix A: Grey literature data sources***

***Government websites***

“soft drinks industry levy” and “soft drink industry levy” and (“sugar tax” and “UK”) and “SDIL” were searched for in the following web pages:

- Association of Directors of Public Health
- Cabinet Office
- Department for Environment, Food & Rural Affairs
- Department of Health and Social Care
- Food Standards Agency
- HM Treasury
- Local Government Association
- NHS England
- Public Health England / Office for Health Improvement and Disparities
- <https://commonslibrary.parliament.uk/>
- <https://lordslibrary.parliament.uk/>
- <https://post.parliament.uk/>

***NGO websites***

“soft drinks industry levy” and “soft drink industry levy” and (“sugar tax” and “UK”) and “SDIL” were searched for in the following web pages:

- Action on Sugar
- Association for the Study of Obesity
- British Heart Foundation
- Cancer Research UK
- Diabetes UK
- Food Foundation
- Jamie Oliver Food Foundation
- The Health Foundation
- The King’s Fund
- Nuffield Foundation
- Obesity Health Alliance
- Obesity UK
- Stroke Association
- Sustain (including Children’s Food Campaign)
- World Cancer Research Fund
- World Obesity Federation
- UK Health Forum

***Industry Body websites***

“soft drinks industry levy” and “soft drink industry levy” and (“sugar tax” and “UK”) and “SDIL” were searched for in the following web pages:

- AB Sugar
- The Association of Convenience Stores
- British Retail Consortium
- British Soft Drinks Association
- Food and Drink Federation
- Food Matters
- Institute of Grocery Distribution
- Institute of Economic Affairs

***Preprint servers***

“soft drinks industry levy” and “soft drink industry levy” and (“sugar tax” and “UK”) and “SDIL” were searched for in the following web pages:

- medrxiv.org/
- bioRxiv.org/

***Ethos***

“soft drinks industry levy” and “soft drink industry levy” and (“sugar tax” and “UK”) and “SDIL”

***Trial Databases***

“soft drinks industry levy” and “soft drink industry levy” and (“sugar tax” and “UK”) and “SDIL” were searched for in the following databases:

- ISCTRN
- PROSPERO

***UK Funders’ websites***

- NIHR
- UKRI sites
- Wellcome Trust

***Targeted searches of google.co.uk***

- In title:”soft drinks industry levy” and “soft drink industry levy” and (“sugar tax” and “UK”) and “SDIL”
- “soft drinks industry levy” and “soft drink industry levy” and “SDIL” filetype:pdf
- “soft drinks industry levy” and “soft drink industry levy” and “SDIL” filetype:pdf site:.uk

**Appendix B: Full database search terms**

***MEDLINE via Ovid***

(tax* OR tariff* OR duty OR duties OR excise* OR levy OR levies OR SDIL).ti,ab,kw. OR taxes/

AND

(drink OR drinks OR beverage* OR fizzy OR sugar* OR sweetened OR soda OR cola OR coke OR carbonated).ti,ab,kw. OR carbonated beverages/ OR cola/

AND

(UK OR “United Kingdom” or “Great Britain" or Engl* OR Wales OR Scotland OR Welsh or Scottish OR “Northern Ireland” OR “Northern Irish”).ti,ab,kw. OR exp United Kingdom/

***Embase via Ovid***

(tax* OR tariff* OR duty OR duties OR excise* OR levy OR levies OR SDIL).ti,ab,kw. OR tax/

AND

(drink OR drinks OR beverage* OR fizzy OR sugar* OR sweetened OR soda OR cola OR coke OR carbonated).ti,ab,kw. OR carbonated beverage/ OR cola/

AND

(UK OR “United Kingdom” or “Great Britain" or Engl* OR Wales OR Scotland OR Welsh or Scottish OR “Northern Ireland” OR “Northern Irish”).ti,ab,kw. OR United Kingdom/

***Web of Science Core Collection***

TS=(tax* OR tariff* OR duty OR duties OR excise* OR levy OR levies OR SDIL)

AND

TS=(drink OR drinks OR beverage* OR fizzy OR sugar* OR sweetened OR soda OR cola OR coke OR carbonated)

AND

TS=(UK OR “United Kingdom” or “Great Britain" or Engl* OR Wales OR Scotland OR Welsh or Scottish OR “Northern Ireland” OR “Northern Irish”)

***Global Health via Ebscohost***

TI (tax* OR tariff* OR duty OR duties OR excise* OR levy OR levies OR SDIL) OR AB (tax* OR tariff* OR duty OR duties OR excise* OR levy OR levies OR SDIL) OR DE “tariffs”

AND

TI (drink OR drinks OR beverage* OR fizzy OR sugar* OR sweetened OR soda OR cola OR coke OR carbonated) OR AB (drink OR drinks OR beverage* OR fizzy OR sugar* OR sweetened OR soda OR cola OR coke OR carbonated) OR DE "soft drinks" OR DE “Cola”

AND

TI (UK OR “United Kingdom” or “Great Britain" or Engl* OR Wales OR Scotland OR Welsh or Scottish OR “Northern Ireland” OR “Northern Irish”) OR AB (UK OR “United Kingdom” or “Great Britain" or Engl* OR Wales OR Scotland OR Welsh or Scottish OR “Northern Ireland” OR “Northern Irish”) OR DE "UK" OR DE "Channel Islands" OR DE "Great Britain" OR DE "Isle of Man" OR DE "Northern Ireland" OR DE “Scotland” OR DE “Wales” OR DE “England”

***EconLit via Ebscohost***

TI (tax* OR tariff* OR duty OR duties OR excise* OR levy OR levies OR SDIL) OR AB (tax* OR tariff* OR duty OR duties OR excise* OR levy OR levies OR SDIL) OR ZU "excise taxes"

AND

TI (drink OR drinks OR beverage* OR fizzy OR sugar* OR sweetened OR soda OR cola OR coke OR carbonated) OR AB (drink OR drinks OR beverage* OR fizzy OR sugar* OR sweetened OR soda OR cola OR coke OR carbonated)

AND

TI (UK OR “United Kingdom” or “Great Britain" or Engl* OR Wales OR Scotland OR Welsh or Scottish OR “Northern Ireland” OR “Northern Irish”) OR AB (UK OR “United Kingdom” or “Great Britain" or Engl* OR Wales OR Scotland OR Welsh or Scottish OR “Northern Ireland” OR “Northern Irish”)

***ERIC via Ebscohost***

TI (tax* OR tariff* OR duty OR duties OR excise* OR levy OR levies OR SDIL) OR AB (tax* OR tariff* OR duty OR duties OR excise* OR levy OR levies OR SDIL) OR KW (tax* OR tariff* OR duty OR duties OR excise* OR levy OR levies OR SDIL) OR DE "Taxes"

AND

TI (drink OR drinks OR beverage* OR fizzy OR sugar* OR sweetened OR soda OR cola OR coke OR carbonated) OR AB (drink OR drinks OR beverage* OR fizzy OR sugar* OR sweetened OR soda OR cola OR coke OR carbonated) OR KW (drink OR drinks OR beverage* OR fizzy OR sugar* OR sweetened OR soda OR cola OR coke OR carbonated)

AND

TI (UK OR “United Kingdom” or “Great Britain" or Engl* OR Wales OR Scotland OR Welsh or Scottish OR “Northern Ireland” OR “Northern Irish”) OR AB (UK OR “United Kingdom” or “Great Britain" or Engl* OR Wales OR Scotland OR Welsh or Scottish OR “Northern Ireland” OR “Northern Irish”) OR KW (UK OR “United Kingdom” or “Great Britain" or Engl* OR Wales OR Scotland OR Welsh or Scottish OR “Northern Ireland” OR “Northern Irish”)

***Sociological Abstracts via ProQuest***

ti(tax* OR tariff* OR duty OR duties OR excise* OR levy OR levies OR SDIL) OR ab(tax* OR tariff* OR duty OR duties OR excise* OR levy OR levies OR SDIL) OR MAINSUBJECT.EXACT("Taxation")

AND

ti(drink OR drinks OR beverage* OR fizzy OR sugar* OR sweetened OR soda OR cola OR coke OR carbonated) OR ab(drink OR drinks OR beverage* OR fizzy OR sugar* OR sweetened OR soda OR cola OR coke OR carbonated)

AND

ti(UK OR “United Kingdom” or “Great Britain" or Engl* OR Wales OR Scotland OR Welsh or Scottish OR “Northern Ireland” OR “Northern Irish”) OR ab(UK OR “United Kingdom” or “Great Britain" or Engl* OR Wales OR Scotland OR Welsh or Scottish OR “Northern Ireland” OR “Northern Irish”)

***PsycInfo via Ebscohost***

TI (tax* OR tariff* OR duty OR duties OR excise* OR levy OR levies OR SDIL) OR AB (tax* OR tariff* OR duty OR duties OR excise* OR levy OR levies OR SDIL) OR KW (tax* OR tariff* OR duty OR duties OR excise* OR levy OR levies OR SDIL)

AND

TI (drink OR drinks OR beverage* OR fizzy OR sugar* OR sweetened OR soda OR cola OR coke OR carbonated) OR AB (drink OR drinks OR beverage* OR fizzy OR sugar* OR sweetened OR soda OR cola OR coke OR carbonated) OR KW (drink OR drinks OR beverage* OR fizzy OR sugar* OR sweetened OR soda OR cola OR coke OR carbonated)

AND

TI (UK OR “United Kingdom” or “Great Britain" or Engl* OR Wales OR Scotland OR Welsh or Scottish OR “Northern Ireland” OR “Northern Irish”) OR AB (UK OR “United Kingdom” or “Great Britain" or Engl* OR Wales OR Scotland OR Welsh or Scottish OR “Northern Ireland” OR “Northern Irish”) OR KW (UK OR “United Kingdom” or “Great Britain" or Engl* OR Wales OR Scotland OR Welsh or Scottish OR “Northern Ireland” OR “Northern Irish”)

***Worldwide Political Science Abstracts via ProQuest***

ti(tax* OR tariff* OR duty OR duties OR excise* OR levy OR levies OR SDIL) OR ab(tax* OR tariff* OR duty OR duties OR excise* OR levy OR levies OR SDIL) OR MAINSUBJECT.EXACT("Taxation")

AND

ti(drink OR drinks OR beverage* OR fizzy OR sugar* OR sweetened OR soda OR cola OR coke OR carbonated) OR ab(drink OR drinks OR beverage* OR fizzy OR sugar* OR sweetened OR soda OR cola OR coke OR carbonated)

AND

ti(UK OR “United Kingdom” or “Great Britain" or Engl* OR Wales OR Scotland OR Welsh or Scottish OR “Northern Ireland” OR “Northern Irish”) OR ab(UK OR “United Kingdom” or “Great Britain" or Engl* OR Wales OR Scotland OR Welsh or Scottish OR “Northern Ireland” OR “Northern Irish”) OR MAINSUBJECT.EXACT("British")

***Appendix C: List of all included studies***

| **First Author, Date ^citation number^** | **Reference** | **Study Methods** |
| --- | --- | --- |
| Allais, 2023^38^ | Allais O, Enderli G, Sassi F, Soler LG. Effective policies to promote sugar reduction in soft drinks: lessons from a comparison of six European countries. European Journal of Public Health. 2023 Dec 1;33(6):1095-101. | Quantitative |
| Bandy, 2020^39^ | Bandy LK, Scarborough P, Harrington RA, Rayner M, Jebb SA. Reductions in sugar sales from soft drinks in the UK from 2015 to 2018. BMC medicine. 2020 Dec;18(1):1-0. | Quantitative |
| Barigozzi, n.d.^40^ | Barigozzi F, Cornelsen L, Mazzocchi M. A Tax is a Signal: Theory and Evidence. | Quantitative |
| Bridge, 2020^41^ | Bridge G, Flint SW, Tench R. An exploration of the portrayal of the UK soft drinks industry levy in UK national newspapers. Public Health Nutrition. 2020 Dec;23(17):3241-9. | Qualitative |
| Buckton, 2018^33^ | Buckton CH, Patterson C, Hyseni L, Katikireddi SV, Lloyd-Williams F, Elliott-Green A, Capewell S, Hilton S. The palatability of sugar-sweetened beverage taxation: a content analysis of newspaper coverage of the UK sugar debate. Plos one. 2018 Dec 5;13(12):e0207576. | Quantitative |
| Chu, 2020^42^ | Chu BT, Irigaray CP, Hillier SE, Clegg ME. The sugar content of children’s and lunchbox beverages sold in the UK before and after the soft drink industry levy. European Journal of Clinical Nutrition. 2020 Apr;74(4):598-603. | Quantitative |
| Cobiac, 2023^43^ | Cobiac LJ, Law C, Smith R, Cummins S, Rutter H, Rayner M, Mytton O, Briggs AD, Tarp Jensen H, Keogh-Brown M, Adams J. Population health and health sector cost impacts of the UK Soft Drinks Industry Levy: a modelling study. medRxiv. 2023:2023-10. [in press, *Public Health Research*, 2025] | Quantitative |
| Cobiac, 2024^44^ | Cobiac, L.J., Rogers, N.T; Adams, J; Cummins, S; Smith, S; Mytton, O; White, M; Scarborough, P. Impact of the UK Soft Drinks Industry Levy on health and health inequalities in children and adolescents in England: an interrupted time series analysis and population health modelling study. Plos Medicine. 2024. 21(3):e1004371. | Quantitative |
| Diez Alonso, 2021^45^ | Díez Alonso D. Essays in public and behavioural economics. (Doctoral dissertation, University of Warwick). | Quantitative |
| Fearne, 2022^46^ | Fearne A, Borzino N, De La Iglesia B, Moffatt P, Robbins M. Using supermarket loyalty card data to measure the differential impact of the UK soft drink sugar tax on buyer behaviour. Journal of Agricultural Economics. 2022 Jun;73(2):321-37. | Quantitative |
| Food Standards Agency, 2020^47^ | Food Standards Agency. Northern Ireland Take Home Food and Drink Purchases (2014 and 2018). 2020. | Quantitative |
| Forde, 2021^48^ | Forde H. Changes to marketing in response to sugary beverage taxation: the soft drinks industry levy in the United Kingdom. 2021. (Doctoral dissertation, University of Cambridge). | Mixed Methods |
| Forde, 2022^49^ | Forde H, Penney TL, White M, Levy L, Greaves F, Adams J. Understanding marketing responses to a tax on sugary drinks: a qualitative interview study in the United Kingdom, 2019. International journal of health policy and management. 2022 Dec;11(11):2618. | Mixed Methods |
| Hashem, 2019^50^ | Hashem KM, He FJ, MacGregor GA. Labelling changes in response to a tax on sugar-sweetened beverages, United Kingdom of Great Britain and Northern Ireland. Bulletin of the World Health Organization. 2019 Dec 12;97(12):818. | Quantitative |
| HM Revenue and Customs, 2021^51^ | HM Revenue and Customs. Soft Drinks Industry Levy statistics commentary 2021. 2021 | Quantitative |
| HM Revenue and Customs, 2022^52^ | HM Revenue and Customs. Soft Drinks Industry Levy statistics commentary 2022. 2022 | Quantitative |
| Huang, 2021^53^ | Huang Y, Theis DR, Burgoine T, Adams J. Trends in energy and nutrient content of menu items served by large UK chain restaurants from 2018 to 2020: an observational study. BMJ Open. 2021 Dec 1;11(12):e054804. | Quantitative |
| Jones, 2023^32^ | Jones CP, Forde H, Penney TL, van Tulleken D, Cummins S, Adams J, Law C, Rutter H, Smith R, White M. Industry views of the UK Soft Drinks Industry Levy: a thematic analysis of elite interviews with food and drink industry professionals, 2018-2020. BMJ Open. 2023 Aug 9;13(8):e072223. | Qualitative |
| Jones, 2024^54^ | Jones CP, Lawlor ER, Forde H, Theis DR, Cummins S, Adams J, Smith R, Rayner M, Rutter H, Penney TL, Alliot O. Parliamentary reaction to the announcement and implementation of the UK Soft Drinks Industry Levy: applied thematic analysis of 2016-2020 parliamentary debates. Public health nutrition. 2024 Jan 24:1-29. | Qualitative |
| Law, 2020a^55^ | Law C, Cornelsen L, Adams J, Penney T, Rutter H, White M, Smith R. An analysis of the stock market reaction to the announcements of the UK Soft Drinks Industry Levy. Economics & Human Biology. 2020 Aug 1;38:100834. | Quantitative |
| Law, 2020b^56^ | Law C, Cornelsen L, Adams J, Pell D, Rutter H, White M, Smith R. The impact of UK soft drinks industry levy on manufacturers’ domestic turnover. Economics & Human Biology. 2020 May 1;37:100866. | Quantitative |
| Luick, 2024^57^ | Luick, M., Bandy, L., Harrington, R., Vijayan, J., Adams, J., Cummins, S., Rayner, M., Rogers, N., Rutter, H., Smith, R., White, N., Scarborough, P. The impact of the UK Soft Drink Industry Levy on the soft drink marketplace, 2017 – 2020: an interrupted time series analysis with comparator series. PLoS One. 2024. 19:6:e0301890 | Quantitative |
| Office for Health Improvement and Disparities, 2022^58^ | Office for Health Improvement and Disparities. Sugar reduction – industry progress 2015 to 2020. 2022. | Quantitative |
| Pell, 2019^59^ | Pell D, Penney T, Hammond D, Vanderlee L, White M, Adams J. Support for, and perceived effectiveness of, the UK soft drinks industry levy among UK adults: cross-sectional analysis of the International Food Policy Study. BMJ open. 2019 Mar 1;9(3):e026698. | Quantitative |
| Tedstone, 2018^60^ | Public Health England. Sugar reduction and wider reformulation programme: Report on progress towards the first 5% reduction and next steps. 2018 | Quantitative |
| Niblett, 2019^61^ | Public Health England. Sugar reduction: Report on progress between 2015 and 2018. 2019 | Quantitative |
| Coyle, 2020^62^ | Public Health England. Sugar reduction: Report on progress between 2015 and 2019. 2020 | Quantitative |
| Rogers, 2020^30^ | Rogers NT, Pell D, Penney TL, Mytton O, Briggs A, Cummins S, Rayner M, Rutter H, Scarborough P, Sharp SJ, Smith RD. Anticipatory changes in British household purchases of soft drinks associated with the announcement of the Soft Drinks Industry Levy: A controlled interrupted time series analysis. PLoS Medicine. 2020 Nov 12;17(11):e1003269. | Quantitative |
| Rogers, 2023a^63^ | Rogers NT, Cummins S, Forde H, Jones CP, Mytton O, Rutter H, Sharp SJ, Theis D, White M, Adams J. Associations between trajectories of obesity prevalence in English primary school children and the UK soft drinks industry levy: An interrupted time series analysis of surveillance data. PLoS Medicine. 2023 Jan 26;20(1):e1004160. | Quantitative |
| Rogers, 2023b^64^ | Rogers NT, Cummins S, Jones CP, Mytton O, Rayner M, Rutter H, White M, Adams J. Estimated changes in free sugar consumption one year after the UK Soft drinks industry levy came into force: controlled interrupted time series analysis of the National Diet and Nutrition Survey (2011-2019). medRxiv. 2023:2023-06.  *(Article published post-analysis: Rogers NT, Cummins S, Jones CP, Mytton O, Rayner M, Rutter H, White M, Adams J. Estimated changes in free sugar consumption one year after the UK soft drinks industry levy came into force: controlled interrupted time series analysis of the National Diet and Nutrition Survey (2011–2019). J Epidemiol Community Health. 2024 Sep 1;78(9):578-84.)* | Quantitative |
| Rogers, 2023c^65^ | Rogers NT, Cummins S, Pell D, Rutter H, Sharp SJ, Smith R, White M, Adams J. Changes in household purchasing of soft drinks following the UK Soft Drinks Industry Levy by household income and composition: controlled interrupted time series analysis, March 2014 to November 2019. medRxiv. 2023 Nov 27:2023-11.  *(Article published post-analysis: Rogers NT, Cummins S, Pell D, Rutter H, Sharp SJ, Smith R, White M, Adams J. Changes in household purchasing of soft drinks following the UK soft drinks industry levy by household income and composition: controlled interrupted time series analysis, March 2014 to November 2019: BMJ Nutrition, Prevention & Health 2025;e000981.)* | Quantitative |
| Rogers, 2023d^66^ | Rogers NT, Conway DI, Mytton O, Roberts CH, Rutter H, Sherriff A, White M, Adams J. Estimated impact of the UK soft drinks industry levy on childhood hospital admissions for carious tooth extractions: interrupted time series analysis. BMJ nutrition, prevention & health. 2023 Dec;6(2):243. | Quantitative |
| Rogers, 2023e^29^ | Rogers, N, Pell D, Mytton O, Penney TL, Briggs A, Cummins S, Jones C, Rayner M, Rutter H, Scarborough P, Sharp SJ, Smith RD. Changes in soft drinks purchased by British households associated with the UK soft drinks industry levy: controlled interrupted time series analysis. BMJ Open 2023; 13:e077059. | Quantitative |
| Rogers, 2023f^67^ | Rogers NT, Cummins S, Jones CP, Mytton OT, Roberts CH, Shaheen SO, Shah SA, Sheikh A, White M, Adams J. The UK Soft Drinks Industry Levy and childhood hospital admissions for asthma in England. medRxiv. 2023 Nov 6:2023-11.  (*Article published post-analysis: Rogers NT, Cummins S, Jones CP, Mytton OT, Roberts CH, Shaheen SO, Shah SA, Sheikh A, White M, Adams J. The UK Soft Drinks Industry Levy and childhood hospital admissions for asthma in England. Nature Communications. 2024 Jun 10;15(1):4934.)* | Quantitative |
| Scarborough, 2020^68^ | Scarborough P, Adhikari V, Harrington RA, Elhussein A, Briggs A, Rayner M, Adams J, Cummins S, Penney T, White M. Impact of the announcement and implementation of the UK Soft Drinks Industry Levy on sugar content, price, product size and number of available soft drinks in the UK, 2015-19: A controlled interrupted time series analysis. PLoS Medicine. 2020 Feb 11;17(2):e1003025. | Quantitative |
| Tarp Jensen, 2025^31^ | Tarp Jensen H, Keogh-Brown MR, Cobiac LJ, Law C, Rogers NT, Rutter H, White M, Smith R. The impact of fiscal food policies on the macro-economy: the case of the UK Soft Drinks Industry Levy. *medRxiv* 2025:2025.04.23.25326252. doi: 10.1101/2025.04.23.25326252. (under review, post-analysis: Health Economics, 2025) | Quantitative |
| Data Study Group team (2022)^69^ | The Alan Turing Institute. Data Study Group Final Report: Sainsbury’s. Investigating the impact of the UK’s Soft Drinks Industry Levy on consumers’ purchases of soft drinks. 2021 | Quantitative |
| Von Philipsborn, 2023^70^ | Von Philipsborn P, Huizinga O, Leibinger A, Rubin D, Burns J, Emmert-Fees K, Pedron S, Laxy M, Rehfuess E. Interim Evaluation of Germany’s Sugar Reduction Strategy for Soft Drinks: Commitments versus Actual Trends in Sugar Content and Sugar Sales from Soft Drinks. Annals of Nutrition and Metabolism. 2023 Sep 21;79(3):282-90. | Quantitative |

***Appendix D: Full list of variables extracted from included studies, and their relationship to nodes identified in the conceptual system map***

| **SDIL conceptual map node** | **New Node** | **Extracted variable** | **First author and date^citation number^** |
| --- | --- | --- | --- |
| Acute and chronic health and wellbeing outcomes | Measured negative health and wellbeing outcomes | Reception boys, obesity prevalence (defined as >95th centile on the UK90 growth charts) | Rogers, 2023a^63^ |
|  |  | Reception girls, obesity prevalence (defined as >95th centile on the UK90 growth charts) | Rogers, 2023a^63^ |
|  |  | Year 6 boys, obesity prevalence (defined as >95th centile on the UK90 growth charts) | Rogers, 2023a^63^ |
|  |  | Year 6 girls, obesity prevalence (defined as >95th centile on the UK90 growth charts) | Rogers, 2023a^63^ |
|  |  | Hospital admissions for carious tooth extractions in all children | Rogers, 2023d^66^ |
|  |  | Overall relative reduction in incidence rates of hospital admissions [asthma] | Rogers, 2023f^67^ |
|  | Predicted negative health and wellbeing outcomes | Cases of cardiovascular diseases | Cobiac, 2023^43^ |
|  |  | Cases of obesity-related cancer | Cobiac, 2023^43^ |
|  |  | Cases of type 2diabetes | Cobiac, 2023^43^ |
|  |  | Current UK population lifetime population health (measured in quality-adjusted life years [QALYs]) | Cobiac, 2023^43^ |
|  |  | Dental caries | Cobiac, 2023^43^ |
|  |  | Prevalence of overweight and obesity in the UK | Cobiac, 2023^43^ |
|  |  | Children and adolescents classified as overweight or obese, in the first ten years after implementation | Cobiac, 2024^44^ |
|  |  | Dental caries in the first ten years after implementation | Cobiac, 2024^43^ |
|  |  | Predicted (positive) impacts on health | Cobiac, 2024^43^ |
|  |  | Predicted impacts on health of children and adolescents in the most deprived areas | Cobiac, 2024^43^ |
|  |  | Predicted impacts on health of children and adolescents in the most deprived areas | Cobiac, 2024^43^ |
|  |  | Future health gains of 86,951 DALYs saved | Tarp Jensen, 2025^31^ |
| Chancellors’ announcement and context (March 2016) | Chancellors’ announcement of the SDIL (16^th^ March 2016) | The announcement (on March 16, 2016) of the two-tiered structure of the UK soft drink levy | Barigozzi, n.d.^40^ |
|  |  | The announcement (on March 16, 2016) of the two-tiered structure of the UK soft drink levy | Barigozzi, n.d.^40^ |
|  |  | The announcement (on March 16, 2016) of the two-tiered structure of the UK soft drink levy | Barigozzi, n.d.^40^ |
|  |  | SDIL announcement | Bridge, 2020^10^ |
|  |  | Month of the announcement of the SDIL | Buckton, 2018^33^ |
|  |  | SDIL announcement | Jones, 2024^54^ |
|  |  | The day of announcement | Law, 2020a^55^ |
|  |  | Announcement of the SDIL | Law, 2020b^56^ |
|  |  | At implementation | Luick, 2024^57^ |
|  |  | At the implementation of the SDIL | Luick, 2024^57^ |
|  |  | At the point of implementation | Luick, 2024^57^ |
|  |  | At the point of implementation of the SDIL (April 2018 | Luick, 2024^57^ |
|  |  | At the point of implementation of the SDIL (April 2018 | Luick, 2024^57^ |
|  |  | At the time of the implementation | Luick, 2024^57^ |
|  |  | 2015 to 2017 | Tedstone, 2018^60^ |
|  |  | 2015 to 2017 | Tedstone, 2018^60^ |
|  |  | 2015 to 2017 | Tedstone, 2018^60^ |
|  |  | Two years post-announcement | Rogers, 2025^65^ |
|  |  | Two years post-announcement | Rogers, 2025^65^ |
|  |  | Two years post-announcement | Rogers, 2025^65^ |
|  |  | Two years post-announcement | Rogers, 2025^65^ |
|  |  | Two years post-announcement | Rogers, 2025^65^ |
|  |  | Two years post-announcement | Rogers, 2025^65^ |
|  |  | Two years post-announcement | Rogers, 2025^65^ |
|  |  | By the implementation of the SDIL | Scarborough, 2020^68^ |
|  |  | By the implementation of the SDIL | Scarborough, 2020^68^ |
|  |  | Two years post-announcement | Rogers, 2025^65^ |
| Health service and social care demand and cost | Predicted health service and social care demand and cost | Monetary benefit for the health system | Cobiac, 2023^43^ |
|  |  | Health-related macroeconomic gains | Tarp Jensen, 2025^31^ |
|  |  | Health-related macroeconomic gains | Tarp Jensen, 2025^31^ |
|  |  | Costs of health care | Cobiac, 2023^43^ |
| Industry levy on SSBs (April 2018) | Implementation of the SDIL (6^th^ April 2018) | SDIL introduction | Bridge, 2020^10^ |
|  |  | Soft Drinks Industry Levy (SDIL) | HM Revenue and Customs, 2021^51^ |
|  |  | Soft Drinks Industry Levy (SDIL) | HM Revenue and Customs, 2022^52^ |
|  |  | From 2018 to 2020 | Huang, 2021^53^ |
|  |  | Post-implementation of SDIL | Law, 2020b^56^ |
|  |  | October 2018 | Luick, 2024^57^ |
|  |  | Between April 2018 and March 2020 | Luick, 2024^57^ |
|  |  | Between April 2018 and March 2020 | Luick, 2024^57^ |
|  |  | Between April 2018 and March 2020 | Luick, 2024^57^ |
|  |  | Between April 2018 and March 2020 | Luick, 2024^57^ |
|  |  | The SDIL | Luick, 2024^57^ |
|  |  | 1 year after implementation | Rogers, 2023e^29^ |
|  |  | 1 year after implementation | Rogers, 2023e^29^ |
| Marketing of SSBs, sugary and other products | Marketing of SSBs, sugary and other products | Changes in marketing expenditure | Forde, 2021^48^ |
|  |  | Marketing decision-making | Forde, 2022^25^ |
| Price of SSBs | Price of SDIL eligible drinks | Price for the high levy category | Luick, 2024^57^ |
|  |  | Price of eligible drinks in the higher levy category | Luick, 2024^57^ |
|  |  | Prices of intervention drinks in the low levy category | Scarborough, 2020^68^ |
|  |  | Price of intervention drinks in the high levy category | Scarborough, 2020^68^ |
|  |  | Consumer prices of UK sugar drinks | Dıez Alonso, 2021^45^ |
|  | Price of SDIL ineligible drinks | Consumer prices of sugar-free alternatives | Dıez Alonso, 2021^45^ |
|  |  | Price for no levy category | Luick, 2024^57^ |
|  |  | Price for no levy category | Luick, 2024^57^ |
|  |  | Prices of drinks in the no levy category | Scarborough, 2020^68^ |
| Public attitudes, awareness, social norms related to SSBs and other products | Attitudes towards SSBs | Negative attitudes to SSBs | Pell, 2019^59^ |
|  | Perceived effectiveness of the SDIL | Perceived effectiveness of the SDIL | Pell, 2019^59^ |
|  |  | Perceived effectiveness of the SDIL | Pell, 2019^59^ |
|  |  | Perceived effectiveness of the SDIL | Pell, 2019^59^ |
|  | Public support for the SDIL | Support for the SDIL | Pell, 2019^59^ |
|  |  | Support for the SDIL | Pell, 2019^59^ |
|  |  | Support for the SDIL | Pell, 2019^59^ |
|  |  | Support for the SDIL | Pell, 2019^59^ |
|  |  | Support for the SDIL | Pell, 2019^59^ |
|  | Social norms to not drinks SSB | Social norms to not drinks SSBs | Pell, 2019^59^ |
|  |  | Social norms to not drinks SSBs | Pell, 2019^59^ |
|  | SSB Health Knowledge | Knowledge of the link between SSBs and obesity | Pell, 2019^59^ |
|  | Trust in health expert messages | Trust in health expert messages | Pell, 2019^59^ |
|  |  | Trusting messages from health experts and the food and beverage industry | Pell, 2019^59^ |
|  | Trusting messages from the food and beverage industry | Trusting messages from the food and beverage industry | Pell, 2019^59^ |
| Public, political and media discourse about sugar and SSBs | Newspaper articles discussing sugar and SSB taxes | Publication of newspaper articles discussing sugar and SSBs taxes | Bridge, 2020^41^ |
|  |  | Publication of newspaper articles discussing sugar and SSBs taxes | Bridge, 2020^41^ |
|  | Oppositional newspaper articles | Oppositional articles outnumbered supportive ones | Buckton, 2018^33^ |
| Purchasing of other products (substitution, complementing, other) | Purchases of alcohol | Purchases of alcohol | Rogers, 2023e^29^ |
|  |  | Substitution to confectionary or alcohol | Rogers, 2023c^30^ |
|  | Purchases of confectionary | Substitution to confectionary or alcohol | Rogers, 2023c^30^ |
|  |  | Purchases of confectionary | Rogers, 2023e^29^ |
| Purchasing of SSBs | Purchases of SDIL eligible drinks | Total volume sales of soft drinks that are subject to the SDIL | Bandy, 2020^39^ |
|  |  | Consumption [purchasing] of soft drinks announced to enter the high tax rate | Barigozzi, n.d.^40^ |
|  |  | Consumption [purchasing] of soft drinks announced to enter the low tax rate | Barigozzi, n.d.^40^ |
|  |  | Total sales of drinks subject to the SDIL | Office for Health Improvement and Disparities, 2022^58^ |
|  |  | Sales of drinks 5-8g of sugar per 100m | Office for Health Improvement and Disparities, 2022^58^ |
|  |  | Sales of drinks over 8g of sugar per 100ml | Office for Health Improvement and Disparities, 2022^58^ |
|  |  | Sales of drinks subject to the levy | Niblett, 2019^61^ |
|  |  | Overall sales (in litres) of drinks subject to the levy | Coyle, 2020^62^ |
|  |  | Volume of purchases in lower levy tier drinks | Rogers, 2023^30^ |
|  |  | Volume purchased in higher levy tier drinks | Rogers, 2023^30^ |
|  |  | Purchased volume of high tier drinks | Rogers, 2023e^29^ |
|  |  | Purchases of low tier drinks | Rogers, 2023e^29^ |
|  | Purchases of SDIL ineligible drinks | Volume sales of low- and zero-sugar | Bandy, 2020^39^ |
|  |  | Consumption [purchasing] of sugar-added soft drinks exempted from the tax | Barigozzi, n.d. ^40^ |
|  |  | Total volume of Diet Soft Drinks | Food Standards Agency, 2020^47^ |
|  |  | Sales of drinks with less than 5g of sugar per 100ml | Office for Health Improvement and Disparities, 2022^58^ |
|  |  | Volume sales of products with levels of sugar below 5g per 100g | Tedstone, 2018^60^ |
|  |  | Volume of household purchases of no levy drinks | Rogers, 2023^30^ |
|  | Purchases of SSBs | Purchase of most popular sugar-sweetened drinks | Fearne, 2022^46^ |
|  |  | Total volume [purchased] of Regular Soft Drinks | Food Standards Agency, 2020^47^ |
|  |  | Total volume of purchases of all drinks combined | Rogers, 2023^30^ |
|  |  | Volumes of soft drinks purchased weekly | Rogers, 2023c^30^ |
|  |  | Volume of all soft drinks purchased | Rogers, 2023e^29^ |
| Reformulation of sugar in existing products (size, shape and content) | Availability of drinks eligible for the SDIL | Proportion in the highest levy category | Luick, 2024^57^ |
|  |  | Proportion in the highest levy category | Luick, 2024^57^ |
|  |  | Number of available drinks that were in the high levy category | Scarborough, 2020^68^ |
|  |  | Proportion of intervention drinks over the lower levy sugar threshold | Scarborough, 2020^68^ |
|  | Availability of drinks not eligible for the SDIL | Proportion of drinks with sugar content below the SDIL levy threshold | Luick, 2024^57^ |
|  |  | Proportion of eligible drinks below the SDIL levy threshold | Luick, 2024^57^ |
|  |  | Number of available control drinks (levy exempt fruit juices and milk-based drinks) | Scarborough, 2020^68^ |
|  | Calorie content of drinks | Mean energy content of the 83 products | Hashem, 2019^50^ |
|  |  | Calories in drinks likely to be consumed on a single occasion | Niblett, 2019^61^ |
|  |  | In the eating out of home sector in the calories for drinks likely to be consumed on a single occasion | Coyle, 2020^62^ |
|  | Calorie content of SDIL eligible drinks | Calorie content of SDIL drinks likely to be consumed on a single occasion | Tedstone, 2018^60^ |
|  |  | Calorie content of drinks subject to the levy likely to be consumed on a single occasion | Niblett, 2019^61^ |
|  | Drinks using non-nutritive sweeteners | Proportion of drinks with non-sugar sweeteners | Luick, 2024^57^ |
|  | Product size of SDIL eligible drinks | [product] volume (ml) of lower levy drinks | Luick, 2024^57^ |
|  |  | [product] volume of products in the higher levy group | Luick, 2024^57^ |
|  |  | Product size of branded high levy and low levy drinks | Scarborough, 2020^68^ |
|  |  | Product size of own-brand high levy drinks | Scarborough, 2020^68^ |
|  |  | Product size of own-brand low levy drinks | Scarborough, 2020^68^ |
|  | Product size of SDIL ineligible drinks | [product] volume (ml) of no levy drinks | Luick, 2024^57^ |
|  | Reformulation of SSBs | Beverages (fruit juice (FJ), juice drinks (JDs) and smoothies (S) specifically targeted at children had reformulated their ingredients | Chu, 2020^42^ |
|  |  | Reformulation | Jones, 2024^54^ |
|  |  | SSB product reformulation | Tarp Jensen, 2025^31^ |
|  |  | Yearly reductions in average sugar content | Allais, 2023^38^ |
|  | Sugar content of drinks | Sales-weighted mean sugar content of soft drinks | Bandy, 2020^39^ |
|  |  | Mean sugar content of the 83 products | Hashem, 2019^50^ |
|  |  | Sugar content in restaurant beverages | Huang, 2021^53^ |
|  |  | Sales weighted average sugar | Office for Health Improvement and Disparities, 2022^58^ |
|  |  | Drinks consumed outside the home simple average total sugar per 100ml | Niblett, 2019^61^ |
|  |  | Amount of sugar in all soft drinks purchased | Rogers, 2023e |
|  |  | Mean sales-weighted sugar content of soft drinks sold | Von Philipsborn, 2023 |
|  | Sugar content of SDIL eligible drinks | Sugar levels per 100ml of own brand and manufacturer branded products for the drinks included in the SDIL | Tedstone, 2018^60^ |
|  |  | Average sugar content of drinks subject to the Soft Drinks Industry Levy (SDIL) | Niblett, 2019^61^ |
|  |  | In the eating out of home sector the simple average total sugar content for drinks subject to the SDIL | Coyle, 2020^62^ |
|  |  | Total sugar content per 100ml for the drinks subject to the levy | Coyle, 2020^62^ |
| Revenue raised | Revenue raised | Total provisional SDIL receipts | HM Revenue and Customs, 2021^51^ |
|  |  | Total provisional SDIL receipts | HM Revenue and Customs, 2022^52^ |
| SSB consumption | SSB consumption | Consumption of SSBs | Pell, 2019^59^ |
| Total sugar consumption | Sugar consumption from drinks | Free sugar consumption from drinks alone | Rogers, 2024^64^ |
|  | Total sugar consumption | Daily consumption of free sugars from the whole diet in children and adults | Rogers, 2024^64^ |
|  |  | Percentage of total dietary energy from free sugars | Rogers, 2024^64^ |
| *N/A – New node* | Food and drink industry preparation for future regulation | Food and drink industry preparation for future regulation in support of health and environmental sustainability | Jones, 2023^32^ |
|  | Food and drink industry profitability, finances and value | Lasting negative effects on drinks industry | Jones, 2023^32^ |
|  |  | Cumulative abnormal returns of soft drink stocks | Law, 2020a^55^ |
|  |  | Stock returns of four UK-operating soft-drink firms listed on the London Stock Exchange | Law, 2020a^55^ |
|  |  | Domestic turnover of UK soft drinks manufacturers | Law, 2020b^56^ |
|  |  | Domestic turnover of UK soft drinks manufacturers | Law, 2020b^56^ |
|  | GDP | Cumulative real GDP | Tarp Jensen, 2025^31^ |
|  | Health Inequality | Slope index of inequality | Cobiac, 2024^44^ |
|  |  | Most vulnerable market segments – families on low incomes | Fearne, 2022^46^ |
|  | Purchases of sugar from all drinks | Volume of sugars sold per capita per day from soft drinks | Bandy, 2020^39^ |
|  |  | Sugar from purchased drinks in England | Cobiac, 2024^44^ |
|  |  | Sugar from purchased drinks in England | Cobiac, 2024^44^ |
|  |  | Sugar from purchased drinks in England | Cobiac, 2024^44^ |
|  |  | Sugar from purchased drinks in England | Cobiac, 2024^44^ |
|  |  | Sugar purchased from all drinks | Rogers, 2023^30^ |
|  |  | High-income households, weekly sugar purchased in soft drinks | Rogers, 2025^65^ |
|  |  | Low-income households, weekly sugar purchased in soft drinks | Rogers, 2025^65^ |
|  |  | Overall purchased weekly sugar in soft drinks | Rogers, 2025^65^ |
|  |  | Quantity of purchased sugar from beverages | Data Study Group team, 2022^69^ |
|  | Purchases of sugar from SDIL eligible drinks | Total sugar purchased per household from drinks subject to the SDIL | Office for Health Improvement and Disparities, 2022^58^ |
|  |  | Total sugar purchased per household from drinks subject to SDIL | Niblett, 2019^61^ |
|  |  | Total sugar purchased per household from drinks subject to the SDIL | Coyle, 2020^62^ |
|  |  | Total sugar sales from the soft drinks subject to the levy | Coyle, 2020^62^ |
|  |  | Sugar purchased in higher levy tier drinks | Rogers, 2025^65^ |
|  |  | Sugar purchased in lower levy tier drinks | Rogers, 2025^65^ |
|  |  | Sugar in purchased high tier drinks | Rogers, 2023e^29^ |
|  |  | Sugar in purchased low tier drinks | Rogers, 2023e^29^ |
|  | Purchases of sugar from SDIL ineligible drinks | Sugar in household purchases of no levy drinks | Rogers, 2025^65^ |
|  | The Soft Drinks Industry Levy (SDIL) | The announcement in 2016 and adoption in 2018 of the UK tax | Allais, 2023^38^ |
|  |  | Between 2015 and 2018 | Bandy, 2020^39^ |
|  |  | Between 2015 and 2018 | Bandy, 2020^39^ |
|  |  | Between 2015 and 2018 | Bandy, 2020^39^ |
|  |  | Between 2015 and 2018 | Bandy, 2020^39^ |
|  |  | In September 2018 | Chu, 2020^42^ |
|  |  | In the first ten years of implementation, the reductions in sugar and overweight/obesity | Cobiac, 2023^43^ |
|  |  | In the first ten years of implementation, the reductions in sugar and overweight/obesity | Cobiac, 2023^43^ |
|  |  | In the first ten years of implementation, the reductions in sugar and overweight/obesity | Cobiac, 2023^43^ |
|  |  | In the first ten years of implementation, the reductions in sugar and overweight/obesity | Cobiac, 2023^43^ |
|  |  | The SDIL | Cobiac, 2023^43^ |
|  |  | The SDIL | Cobiac, 2023^43^ |
|  |  | The SDIL | Cobiac, 2023^43^ |
|  |  | The SDIL | Cobiac, 2023^43^ |
|  |  | Effects of SDIL | Cobiac, 2024^44^ |
|  |  | The SDIL | Cobiac, 2024^44^ |
|  |  | The SDIL | Dıez Alonso, 2021^45^ |
|  |  | The SDIL | Dıez Alonso, 2021^45^ |
|  |  | Impact of the sugar tax | Fearne, 2022^46^ |
|  |  | Impact of the sugar tax | Fearne, 2022^46^ |
|  |  | Introduction of the Soft Drinks Industry Levy | Food Standards Agency, 2020^47^ |
|  |  | Introduction of the Soft Drinks Industry Levy | Food Standards Agency, 2020^47^ |
|  |  | The announcement and implementation of the SDIL. | Forde, 2021^48^ |
|  |  | The SDIL | Forde, 2022^49^ |
|  |  | Between 2014 and 2018 | Hashem, 2019^50^ |
|  |  | Between 2014 and 2018 | Hashem, 2019^50^ |
|  |  | The SDIL | Jones, 2023^32^ |
|  |  | The SDIL | Jones, 2023^32^ |
|  |  | Beyond the day of the SDIL announcement | Law, 2020a^55^ |
|  |  | From 2015 | Office for Health Improvement and Disparities, 2022^58^ |
|  |  | From 2015 | Office for Health Improvement and Disparities, 2022^58^ |
|  |  | Between 2015 and 2020 | Office for Health Improvement and Disparities, 2022^58^ |
|  |  | Between 2015 and 2020 | Office for Health Improvement and Disparities, 2022^58^ |
|  |  | Between 2015 and 2020 | Office for Health Improvement and Disparities, 2022^58^ |
|  |  | Between 2015 and 2020 | Office for Health Improvement and Disparities, 2022^58^ |
|  |  | Between 2015 and 2018 | Niblett, 2019^61^ |
|  |  | Between 2015 and 2018 | Niblett, 2019^61^ |
|  |  | Between 2015 and 2018 | Niblett, 2019^61^ |
|  |  | Between 2015 and 2018 | Niblett, 2019^61^ |
|  |  | Between 2015 and 2018 | Niblett, 2019^61^ |
|  |  | Between 2015 and 2018 | Niblett, 2019^61^ |
|  |  | Between 2015 and 2019 | Coyle, 2020^62^ |
|  |  | Between 2015 and 2019 | Coyle, 2020^62^ |
|  |  | Between 2015 and 2019 | Coyle, 2020^62^ |
|  |  | Between 2015 and 2019 | Coyle, 2020^62^ |
|  |  | Between 2015 and 2019 | Coyle, 2020^62^ |
|  |  | Between 2015 and 2019 | Coyle, 2020^62^ |
|  |  | September 2013 to November 2019 | Rogers, 2023a^63^ |
|  |  | September 2013 to November 2019 | Rogers, 2023a^63^ |
|  |  | September 2013 to November 2019 | Rogers, 2023a^63^ |
|  |  | September 2013 to November 2019 | Rogers, 2023a^63^ |
|  |  | SDIL | Rogers, 2023b^64^ |
|  |  | SDIL | Rogers, 2023b^64^ |
|  |  | SDIL | Rogers, 2023b^64^ |
|  |  | Between March 2014 to November 2019 | Rogers, 2023c^65^ |
|  |  | Between March 2014 to November 2019 | Rogers, 2023c^65^ |
|  |  | Between March 2014 to November 2019 | Rogers, 2023c^65^ |
|  |  | Between March 2014 to November 2019 | Rogers, 2023c^65^ |
|  |  | Between March 2014 to November 2019 | Rogers, 2023c^65^ |
|  |  | SDIL | Rogers, 2023d^66^ |
|  |  | In March 2019 | Rogers, 2023e^29^ |
|  |  | In March 2019 | Rogers, 2023e^29^ |
|  |  | In March 2019 | Rogers, 2023e^29^ |
|  |  | In March 2019 | Rogers, 2023e^29^ |
|  |  | In March 2019 | Rogers, 2023e^29^ |
|  |  | In March 2019 | Rogers, 2023e^29^ |
|  |  | 22 months post-SDIL | Rogers, 2023f^67^ |
|  |  | After implementation of the SDIL | Scarborough, 2020^68^ |
|  |  | After implementation of the SDIL | Scarborough, 2020^68^ |
|  |  | After implementation of the SDIL | Scarborough, 2020^68^ |
|  |  | In February 2019 | Scarborough, 2020^68^ |
|  |  | In February 2019 | Scarborough, 2020^68^ |
|  |  | Between September 2015 and February 2019 | Scarborough, 2020^68^ |
|  |  | In February 2019 | Scarborough, 2020^68^ |
|  |  | The SDIL | Tarp Jensen, 2025^31^ |
|  |  | The SDIL | Tarp Jensen, 2025^31^ |
|  |  | The SDIL | Tarp Jensen, 2025^31^ |
|  |  | 2016-2019 | Data Study Group team, 2021^69^ |
|  |  | United Kingdom, which adopted a soft drinks tax in 2017 | Von Philipsborn, 2023^70^ |

***Appendix E: Connections extracted from included studies, by frequency of connection***

| **No. of studies** | **Origin Node** | **Polarity of connection** | **Destination Node** | **Studies containing connections (First author, date)** |
| --- | --- | --- | --- | --- |
| 8 | The Soft Drinks Industry Levy (SDIL) | Negative | Purchases of SDIL eligible drinks | Bandy, 2020a; Barigozzi, n.d.; Barigozzi, n.d.; Office for Health Improvement and Disparities, 2022; Office for Health Improvement and Disparities, 2022; Rogers, 2025; Rogers, 2023e; Rogers, 2023e^29, 39, 40, 58, 65^ |
| 8 | The Soft Drinks Industry Levy (SDIL) | Negative | Sugar content of drinks | Bandy, 2020a; Allais, 2023; Hashem, 2019; Huang, 2021; Office for Health Improvement and Disparities, 2022; Niblett, 2019; Rogers, 2023e; Von Philipsborn, 2023^29, 38, 39, 50, 53, 58, 61, 70^ |
| 7 | The Soft Drinks Industry Levy (SDIL) | Negative | Predicted negative health and wellbeing outcomes | Cobiac, 2023; Cobiac, 2023; Cobiac, 2023; Cobiac, 2023; Cobiac, 2023; Cobiac, 2023; Tarp Jensen, 2025^31, 43^ |
| 7 | The Soft Drinks Industry Levy (SDIL) | Negative | Purchases of sugar from SDIL eligible drinks | Rogers, 2025; Office for Health Improvement and Disparities, 2022; Niblett, 2019; Coyle, 2020; Coyle, 2020; Rogers, 2023e; Rogers, 2023e^29, 58, 61, 62, 65^ |
| 5 | The Soft Drinks Industry Levy (SDIL) | Negative | Purchases of sugar from all drinks | Bandy, 2020a; Cobiac, 2024; Rogers, 2023c; Rogers, 2023c; Data Study Group team, 2021^39, 44, 66, 69^ |
| 5 | The Soft Drinks Industry Levy (SDIL) | Positive | Purchases of SDIL ineligible drinks | Bandy, 2020a; Food Standards Agency, 2020 ; Office for Health Improvement and Disparities, 2022; Tedstone, 2018; Rogers, 2025^39, 47, 58, 60, 65^ |
| 4 | The Soft Drinks Industry Levy (SDIL) | Negative | Availability of drinks eligible for the SDIL | Luick, 2024.; Luick, 2024..; Scarborough, 2020; Scarborough, 2020^57, 68^ |
| 4 | The Soft Drinks Industry Levy (SDIL) | Positive | Purchases of SDIL eligible drinks | Rogers, 2025; Office for Health Improvement and Disparities, 2022; Niblett, 2019; Coyle, 2020^58, 61, 62, 65^ |
| 4 | The Soft Drinks Industry Levy (SDIL) | Negative | Sugar content of SDIL eligible drinks | Tedstone, 2018; Niblett, 2019; Coyle, 2020; Coyle, 2020^60-62^ |
| 3 | Purchases of sugar from all drinks | Negative | Predicted negative health and wellbeing outcomes | Cobiac, 2024; Cobiac, 2024; Cobiac, 2024^44^ |
| 3 | The Soft Drinks Industry Levy (SDIL) | Negative | Measured negative health and wellbeing outcomes | Rogers, 2023a; Rogers, 2023d; Rogers, 2023f^63, 66, 67^ |
| 3 | The Soft Drinks Industry Levy (SDIL) | Unclear | Measured negative health and wellbeing outcomes | Rogers, 2023a; Rogers, 2023a; Rogers, 2023a^63^ |
| 3 | The Soft Drinks Industry Levy (SDIL) | Negative | Calorie content of drinks | Hashem, 2019; Niblett, 2019; Coyle, 2020^50, 61, 62^ |
| 3 | The Soft Drinks Industry Levy (SDIL) | Unclear | Food and drink industry profitability, finances and value | Jones, 2023; Law, 2020a; Law, 2020b^32, 55, 56^ |
| 3 | The Soft Drinks Industry Levy (SDIL) | Negative | Predicted health service and social care demand and cost | Cobiac, 2023; Cobiac, 2023; Jensen, 2025^31, 43^ |
| 3 | The Soft Drinks Industry Levy (SDIL) | Positive | Price of SDIL eligible drinks | Luick, 2024; Dıez Alonso, 2021; Scarborough, 2020^45, 57, 68^ |
| 3 | The Soft Drinks Industry Levy (SDIL) | Positive | Price of SDIL ineligible drinks | Luick, 2024.; Dıez Alonso, 2021; Scarborough, 2020^45, 57, 68^ |
| 3 | The Soft Drinks Industry Levy (SDIL) | Unclear | Product size of SDIL eligible drinks | Luick, 2024; Scarborough, 2020^57, 68^ |
| 2 | The Soft Drinks Industry Levy (SDIL) | Positive | Availability of drinks not eligible for the SDIL | Luick, 2024; Luick, 2024^57^ |
| 2 | The Soft Drinks Industry Levy (SDIL) | Negative | Calorie content of SDIL eligible drinks | Tedstone, 2018; Public Health Niblett, 2019^60, 61^ |
| 2 | The Soft Drinks Industry Levy (SDIL) | Negative | Food and drink industry profitability, finances and value | Law, 2020a; Law, 2020b^55, 56^ |
| 2 | The Soft Drinks Industry Levy (SDIL) | Negative | Health Inequality | Cobiac, 2024; Fearne, 2022^44, 46^ |
| 2 | The Soft Drinks Industry Levy (SDIL) | Positive | Newspaper articles discussing sugar and SSB taxes | Bridge, 2020; Bridge, 2020^41^ |
| 2 | The Soft Drinks Industry Levy (SDIL) | Negative | Product size of SDIL eligible drinks | Luick, 2024; Scarborough, 2020^57, 68^ |
| 2 | The Soft Drinks Industry Levy (SDIL) | Unclear | Purchases of alcohol | Rogers, 2023c; Rogers, 2023e^29, 65^ |
| 2 | The Soft Drinks Industry Levy (SDIL) | Unclear | Purchases of confectionary | Rogers, 2023e; Rogers, 2023c^29, 65^ |
| 2 | The Soft Drinks Industry Levy (SDIL) | Negative | Purchases of SSBs | Fearne, 2022; Food Standards Agency, 2020^46, 47^ |
| 2 | The Soft Drinks Industry Levy (SDIL) | Unclear | Purchases of SSBs | Rogers, 2025; Rogers, 2023c^65^ |
| 2 | The Soft Drinks Industry Levy (SDIL) | Positive | Purchases of sugar from all drinks | Rogers, 2025; Rogers, 2023c^65^ |
| 2 | The Soft Drinks Industry Levy (SDIL) | Positive | Reformulation of SSBs | Jones, 2024; Chu, 2020^42, 54^ |
| 2 | The Soft Drinks Industry Levy (SDIL) | Positive | Revenue raised | HM Revenue and Customs, 2021; HM Revenue and Customs, 2022^51, 52^ |
| 1 | Attitudes towards SSBs | Positive | Perceived effectiveness of the SDIL | Pell, 2019^59^ |
| 1 | Predicted negative health and wellbeing outcomes | Negative | Predicted negative health and wellbeing outcomes | Cobiac, 2024^44^ |
| 1 | Reformulation of SSBs | Negative | Predicted health service and social care demand and cost | Tarp Jensen, 2025^31^ |
| 1 | SSB Consumption | Negative | Public support for the SDIL | Pell, 2019^59^ |
| 1 | SSB Health Knowledge | Positive | Public support for the SDIL | Pell, 2019^59^ |
| 1 | The Soft Drinks Industry Levy (SDIL) | Unclear | Availability of drinks not eligible for the SDIL | Scarborough, 2020^68^ |
| 1 | Social norms to not drinks SSBs | Positive | Perceived effectiveness of the SDIL | Pell, 2019^59^ |
| 1 | Social norms to not drinks SSBs | Positive | Public support for the SDIL | Pell, 2019^59^ |
| 1 | The Soft Drinks Industry Levy (SDIL) | Positive | Drinks using non-nutritive sweeteners | Luick, 2024^57^ |
| 1 | The Soft Drinks Industry Levy (SDIL) | Positive | Food and drink industry preparation for future regulation | Jones, 2023^32^ |
| 1 | The Soft Drinks Industry Levy (SDIL) | Negative | GDP | Tarp Jensen, 2025^31^ |
| 1 | The Soft Drinks Industry Levy (SDIL) | Positive | Marketing of SSBs, sugary and other products | Forde, 2022^49^ |
| 1 | The Soft Drinks Industry Levy (SDIL) | Unclear | Marketing of SSBs, sugary and other products | Forde, 2021^48^ |
| 1 | The Soft Drinks Industry Levy (SDIL) | Positive | Oppositional newspaper articles | Buckton, 2018^33^ |
| 1 | The Soft Drinks Industry Levy (SDIL) | Negative | Price of SDIL eligible drinks | Scarborough, 2020^68^ |
| 1 | The Soft Drinks Industry Levy (SDIL) | Unclear | Price of SDIL eligible drinks | Luick, 2024^57^ |
| 1 | The Soft Drinks Industry Levy (SDIL) | Negative | Price of SDIL ineligible drinks | Luick, 2024^57^ |
| 1 | The Soft Drinks Industry Levy (SDIL) | Positive | Product size of SDIL eligible drinks | Scarborough, 2020^68^ |
| 1 | The Soft Drinks Industry Levy (SDIL) | Unclear | Purchases of SDIL ineligible drinks | Barigozzi, n.d.^40^ |
| 1 | The Soft Drinks Industry Levy (SDIL) | Positive | Purchases of SSBs | Rogers, 2023e^29^ |
| 1 | The Soft Drinks Industry Levy (SDIL) | Positive | Purchases of sugar from SDIL eligible drinks | Rogers, 2025^65^ |
| 1 | The Soft Drinks Industry Levy (SDIL) | Positive | Purchases of sugar from SDIL ineligible drinks | Rogers, 2025^65^ |
| 1 | The Soft Drinks Industry Levy (SDIL) | Negative | Sugar consumption from drinks | Rogers, 2023b^64^ |
| 1 | The Soft Drinks Industry Levy (SDIL) | Negative | Total sugar consumption | Rogers, 2023b^64^ |
| 1 | The Soft Drinks Industry Levy (SDIL) | Unclear | Total sugar consumption | Rogers, 2023b^64^ |
| 1 | Trust in health expert messages | Positive | Perceived effectiveness of the SDIL | Pell, 2019^59^ |
| 1 | Trust in health expert messages | Positive | Public support for the SDIL | Pell, 2019^59^ |
| 1 | Trusting messages from the food and beverage industry | Positive | Perceived effectiveness of the SDIL | Pell, 2019^59^ |
| 1 | Trusting messages from the food and beverage industry | Negative | Public support for the SDIL | Pell, 2019^59^ |

***Appendix F: Consistency of connections****

| **Origin Node** | **Destination Node** | **Destination Node Group** | **Number of Connections** | | | | **% Connections with the same polarity** |
| --- | --- | --- | --- | --- | --- | --- | --- |
|  |  |  | **Total** | **Positive polarity** | **Neutral polarity** | **Negative polarity** |  |
| The SDIL | Sugar content of drinks | Market characteristics of soft drinks | 8 | 0 | 0 | 8 | 100 |
| The SDIL | Purchases of sugar from SDIL eligible drinks | Purchasing of soft drinks | 8 | 1 | 0 | 7 | 88 |
| The SDIL | Predicted negative health and wellbeing outcomes | Health related outcomes | 7 | 0 | 0 | 7 | 100 |
| The SDIL | Purchases of SDIL ineligible drinks | Purchasing of soft drinks | 6 | 5 | 1 | 0 | 83 |
| The SDIL | Availability of drinks eligible for the SDIL | Market characteristics of soft drinks | 4 | 0 | 0 | 4 | 100 |
| The SDIL | Sugar content of SDIL eligible drinks | Market characteristics of soft drinks | 4 | 0 | 0 | 4 | 100 |
| The SDIL | Price of SDIL ineligible drinks | Market characteristics of soft drinks | 4 | 3 | 0 | 1 | 75 |
| Purchases of sugar from all drinks | Predicted negative health and wellbeing outcomes | Health related outcomes | 3 | 0 | 0 | 3 | 100 |
| The SDIL | Calorie content of drinks | Market characteristics of soft drinks | 3 | 0 | 0 | 3 | 100 |
| The SDIL | Predicted health service and social care demand and cost | Industry & economy | 3 | 0 | 0 | 3 | 100 |
| The SDIL | Calorie content of SDIL eligible drinks | Market characteristics of soft drinks | 2 | 0 | 0 | 2 | 100 |
| The SDIL | Health Inequality | Health related outcomes | 2 | 0 | 0 | 2 | 100 |
| The SDIL | Newspaper articles discussing sugar and SSB taxes | Media discourse | 2 | 2 | 0 | 0 | 100 |
| The SDIL | Purchases of alcohol | Purchasing of other products | 2 | 0 | 2 | 0 | 100 |
| The SDIL | Purchases of confectionary | Purchasing of other products | 2 | 0 | 2 | 0 | 100 |
| The SDIL | Reformulation of SSBs | Market characteristics of soft drinks | 2 | 2 | 0 | 0 | 100 |
| The SDIL | Revenue raised | Industry & economy | 2 | 2 | 0 | 0 | 100 |
| The SDIL | Purchases of SDIL eligible drinks | Purchasing of soft drinks | 12 | 4 | 0 | 8 | 67 |
| The SDIL | Purchases of sugar from all drinks | Purchasing of soft drinks | 7 | 2 | 0 | 5 | 71 |
| The SDIL | Measured negative health and wellbeing outcomes | Health related outcomes | 6 | 0 | 3 | 3 | 50 |
| The SDIL | Product size of SDIL eligible drinks | Market characteristics of soft drinks | 6 | 1 | 3 | 2 | 50 |
| The SDIL | Food and drink industry profitability, finances and value | Industry & economy | 5 | 0 | 3 | 2 | 60 |
| The SDIL | Price of SDIL eligible drinks | Market characteristics of soft drinks | 5 | 3 | 1 | 1 | 60 |
| The SDIL | Purchases of SSBs | Purchasing of soft drinks | 5 | 1 | 2 | 2 | 40 |
| The SDIL | Availability of drinks not eligible for the SDIL | Market characteristics of soft drinks | 3 | 2 | 1 | 0 | 67 |
| The SDIL | Total sugar consumption | Sugar consumption | 2 | 0 | 1 | 1 | 50 |
| The SDIL | Marketing of SSBs, sugary and other products | Industry & economy | 2 | 1 | 1 | 0 | 50 |
| Attitudes towards SSBs | Perceived effectiveness of the SDIL | Public attitudes, awareness | 1 | 1 | 0 | 0 | 100 |
| Reformulation of SSBs | Predicted health service and social care demand and cost | Industry & economy | 1 | 0 | 0 | 1 | 100 |
| Social norms to not drinks SSBs | Perceived effectiveness of the SDIL | Public attitudes, awareness | 1 | 1 | 0 | 0 | 100 |
| Social norms to not drinks SSBs | Public support for the SDIL | Public attitudes, awareness | 1 | 1 | 0 | 0 | 100 |
| SSB Consumption | Public support for the SDIL | Public attitudes, awareness | 1 | 0 | 0 | 1 | 100 |
| SSB Health Knowledge | Public support for the SDIL | Public attitudes, awareness | 1 | 1 | 0 | 0 | 100 |
| The SDIL | Drinks using non-nutritive sweeteners | Market characteristics of soft drinks | 1 | 1 | 0 | 0 | 100 |
| The SDIL | Food and drink industry preparation for future regulation | Industry & economy | 1 | 1 | 0 | 0 | 100 |
| The SDIL | GDP | Industry & economy | 1 | 0 | 0 | 1 | 100 |
| The SDIL | Oppositional newspaper articles | Media discourse | 1 | 1 | 0 | 0 | 100 |
| The SDIL | Purchases of sugar from SDIL ineligible drinks | Purchasing of soft drinks | 1 | 1 | 0 | 0 | 100 |
| The SDIL | Sugar consumption from drinks | Sugar consumption | 1 | 0 | 0 | 1 | 100 |
| Trust in health expert messages | Perceived effectiveness of the SDIL | Public attitudes, awareness | 1 | 1 | 0 | 0 | 100 |
| Trust in health expert messages | Public support for the SDIL | Public attitudes, awareness | 1 | 1 | 0 | 0 | 100 |
| Trusting messages from the food & beverage industry | Perceived effectiveness of the SDIL | Public attitudes, awareness | 1 | 1 | 0 | 0 | 100 |
| Trusting messages from the food & beverage industry | Public support for the SDIL | Public attitudes, awareness | 1 | 0 | 0 | 1 | 100 |

**Highest % of connections with the same polarity. Green shaded cells = ‘high consistency’ (where there are 2 or more studies reporting the same connection and >75% have the same polarity); Yellow shaded cells – ‘low consistency’ (where there are 2 or more studies reporting the same connection and <75% have the same polarity); Blue shaded cells = connections only reported by one study.*
